# A DNA methylome biosignature in alveolar macrophages from TB-exposed individuals predicts exposure to mycobacteria

**DOI:** 10.1101/2021.03.16.21253732

**Authors:** Jyotirmoy Das, Nina Idh, Isabelle Pehrson, Jakob Paues, Maria Lerm

## Abstract

Several studies have identified biomarkers for tuberculosis (TB) diagnosis based on blood cell transcriptomics. Here, we instead studied epigenomics in the lung compartment by obtaining induced sputum from subjects included in a TB contact tracing. CD3- and HLA-DR-positive cells were isolated from the collected sputum and DNA methylome analyses performed. Unsupervised cluster analysis revealed that DNA methylomes of cells from TB-exposed individuals and controls appeared as separate clusters and the numerous genes that were differentially methylated were functionally connected. The enriched pathways were strongly correlated to data from published work on protective heterologous immunity to TB. Taken together, our results demonstrate that epigenetic changes related to trained immunity occurs in the pulmonary immune cells of TB-exposed individuals and that a DNA methylation signature can be derived from the DNA methylome. Such a signature can be further developed for clinical use as a marker of TB exposure.

## Introduction

Tuberculosis (TB) is caused by *Mycobacterium tuberculosis,* which is transmitted between individuals through the inhalation of aerosols generated by coughing ^1^. The disease still claims more than one million human lives annually and an expansion of the current toolkit for diagnosis, prevention and treatment is critical for reaching the United Nations’ Sustainable Development Goals for 2030 of ending the TB epidemic ^2^. In response to the urgent need for new approaches to diagnose TB, recent studies identified TB-specific biosignatures based on RNA transcription profiles in peripheral blood of TB-infected individuals ^3, 4^. Similar TB biosignatures have been identified that could predict disease progression ^5, 6^ and reflect treatment monitoring ^6, 7^. Biosignatures for the detection of TB-exposure in easily accessible clinical samples could provide a basis for the development of novel effective point-of-care diagnostic tools.

The only available TB vaccine is Bacillus Calmette Guérin (BCG), which is based on live attenuated *M. bovis* ^8^. In a recent study, we showed that administration of the BCG vaccine to healthy subjects induced profound epigenetic alterations in immune cells, which correlated with enhanced anti-mycobacterial activity in macrophages isolated from the vaccinees^9^. The changes were reflected in the DNA methylome, with the strongest response being recorded within weeks after vaccination^9^. Our observation that BCG induces alterations of the DNA methylome of immune cells has later been confirmed by others ^10, 11^. Since BCG vaccination reflects an *in vivo* interaction between immune cells and viable mycobacteria, we hypothesized that natural exposure to *M. tuberculosis* would induce similar changes not only in TB patients, but also in individuals who have been exposed to TB. To enable recruitment of control subjects with very low likelihood of previous TB exposure, we performed the study in a low-endemic setting. We established a protocol for the isolation of distinct cell populations from induced sputum^12^, from which DNA could be isolated. Analyses of DNA methylomes of immune cells isolated from lungs and peripheral blood allowed us to identify distinct DNA methylation signatures in TB-exposed individuals. The signature was most prominent in the lung-derived cell populations. Pathway analyses revealed strong overlaps with previous studies on BCG-induced epigenetic signatures that could be correlated with protection against *M. tuberculosis*. We also identified pathway overlap with previous work on trained immunity induced by β-glucan from *Candida albicans*. In conclusion, we found a distinct pattern of DNA methylome changes in immune cells isolated from lungs in individuals with documented exposure to TB. The alterations strongly overlap with pathways described as reflecting trained immunity and an enhanced anti-mycobacterial response. The identified signature has potential to be used as a tool to identify TB exposure in low-endemic settings.

## Results

### Study design

To determine epigenetic changes in the immune cells in TB-exposed individuals, we recruited subjects enrolled in a routinely performed TB contact tracing at Linköping University Hospital, Sweden. Age-matched individuals were included as controls (Table 1). The index case was diagnosed with drug-sensitive pulmonary TB and had completed two out of six months of standard treatment at the time of sample collection. All included subjects except one (a TB contact) were BCG-vaccinated (Table 1). Interferon-Gamma Release Assay (IGRA) status was determined and among the exposed individuals, two were positive (including the index case) and among the controls, one individual (C2) was classified as ‘borderline’-positive ^13^ (Table 1). From induced sputum, HLA-DR-positive (antigen-presenting cells, dominated by macrophages ^14^ and CD3-positive (T cell) populations were purified, whereas the PBMC fraction extracted from blood were kept as a mixed population (Fig. 1).

**Figure 1:**
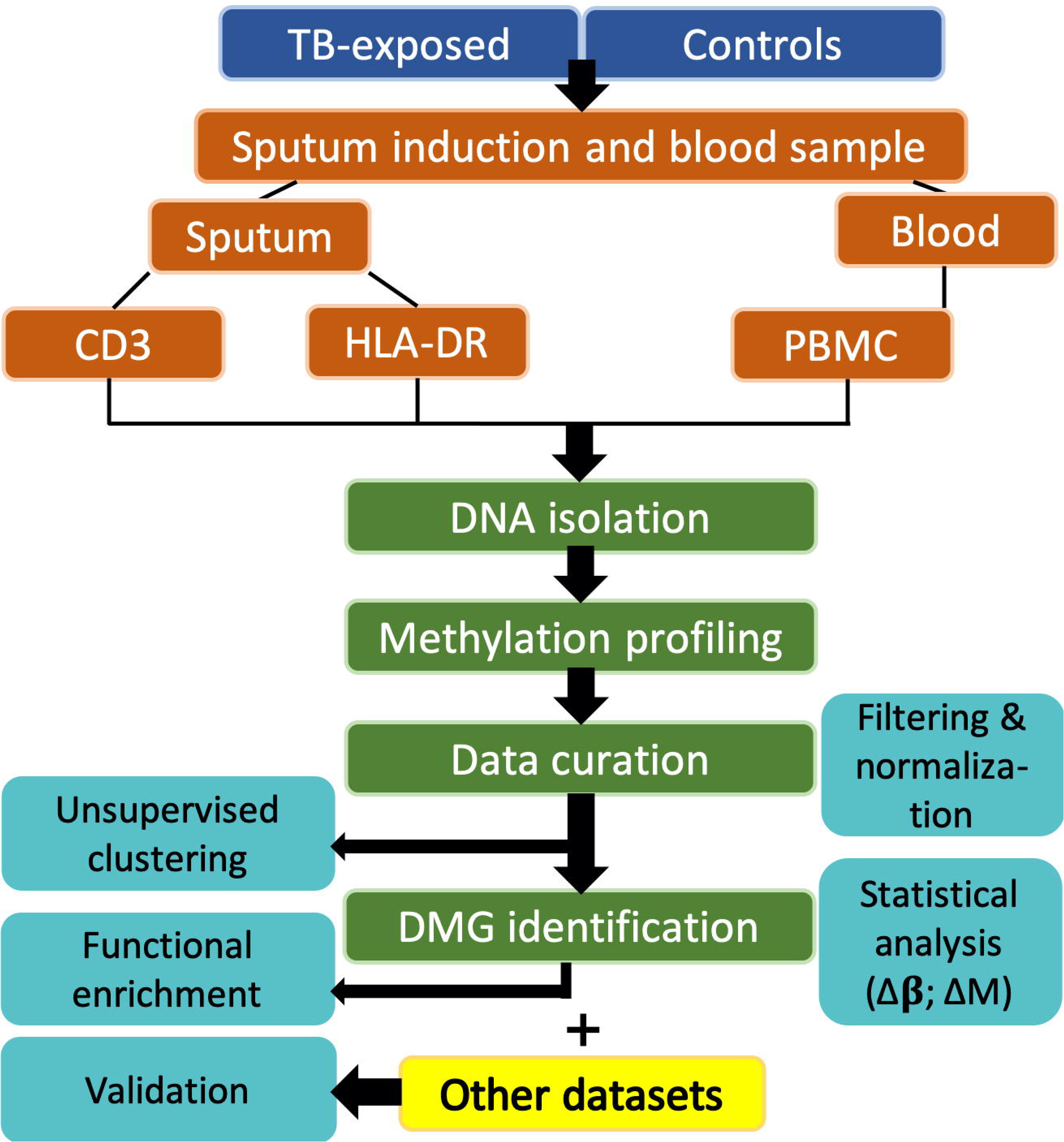
Flow chart of the project workflow.

**Table 1:** Demographic data of the participants.

| Characteristics | Exposed (n = 4) | Controls (n = 6) |
| --- | --- | --- |
| Mean age (year)<sup>†</sup> | 36±12 | 27±6 |
| Mean height (cm)<sup>†</sup> | 173±4 | 176±7 |
| Mean weight (kg)<sup>†</sup> | 69±8 | 77±16 |
| Mean Body Mass Index (BMI)<sup>†</sup> | 23±3 | 24±5 |
| Sex (male/female) | 1/3 | 4/2 |
| Smoking (current/previous/never) | 0/1/3 | 0/0/6 |
| BCG (yes/no) | 3/1 | 6/0 |
| IGRA-positive/IGRA-negative | 2/2 | 1*/5 |

<sup>†</sup>The standard deviation of the mean values is added to the age, height, weight and BMI.
\* Borderline-positive.

### DNA methylome data from TB-exposed individuals form a separate cluster

DNA isolation from the studied cell populations was followed by global DNA methylation analysis using the Illumina 450K protocol. After curation of the data^15^, the datasets were subjected to unsupervised hierarchical cluster analysis based on DNA CpG methylation β-values (see project work-flow, Fig. 1). This approach accurately clustered the participants into TB-exposed and controls based on the DNA methylome data derived from both HLA-DR- and CD3-positive cell populations. (Fig. 2a, b). On the other hand, in the PBMC-derived dataset, the TB index case appeared outside the clusters and two of the controls clustered with the other exposed individuals, one of them (“Con_2”) being the individual identified as border line-positive in the IGRA test (Fig. 2c).

**Figure 2:**
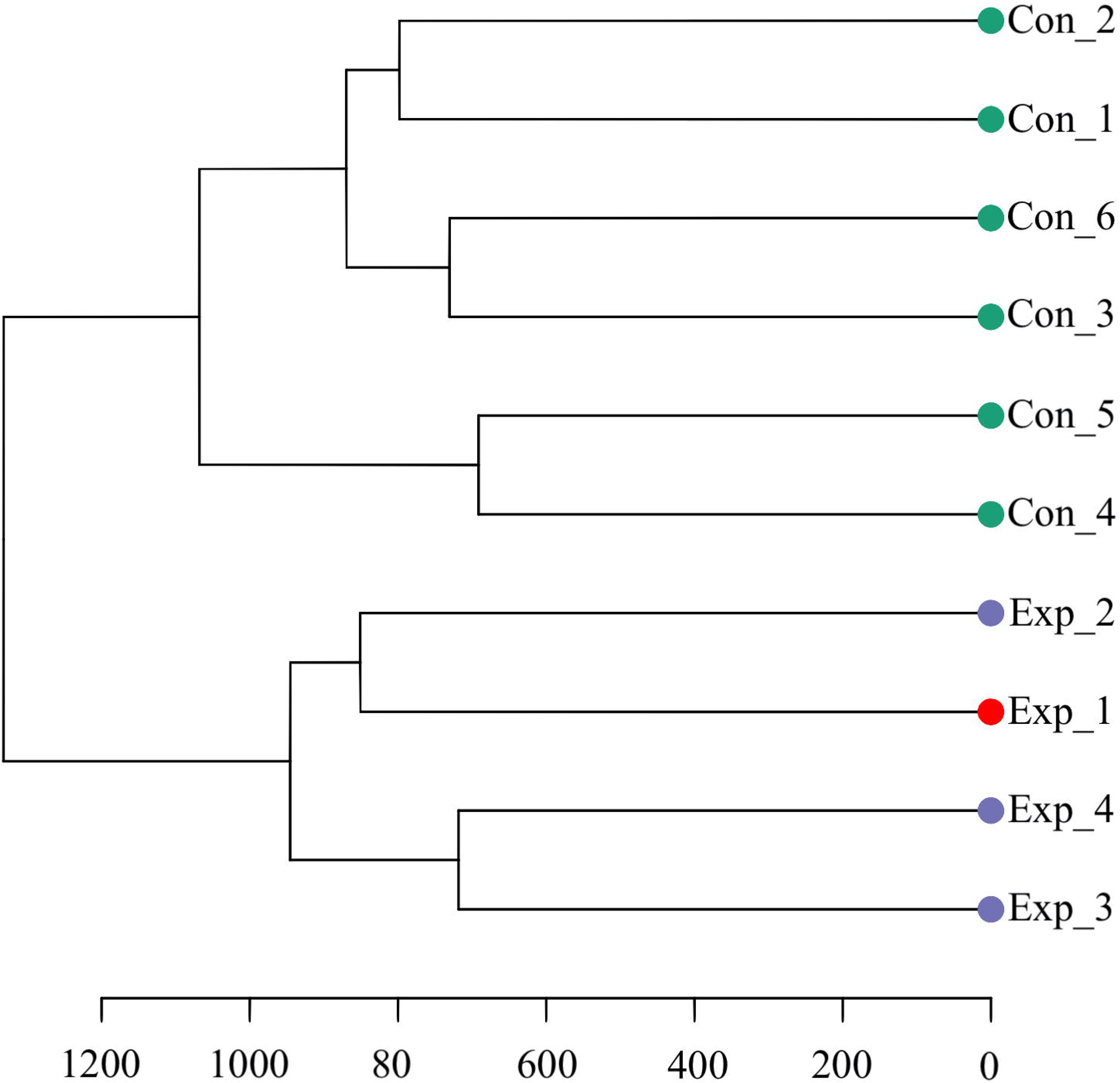
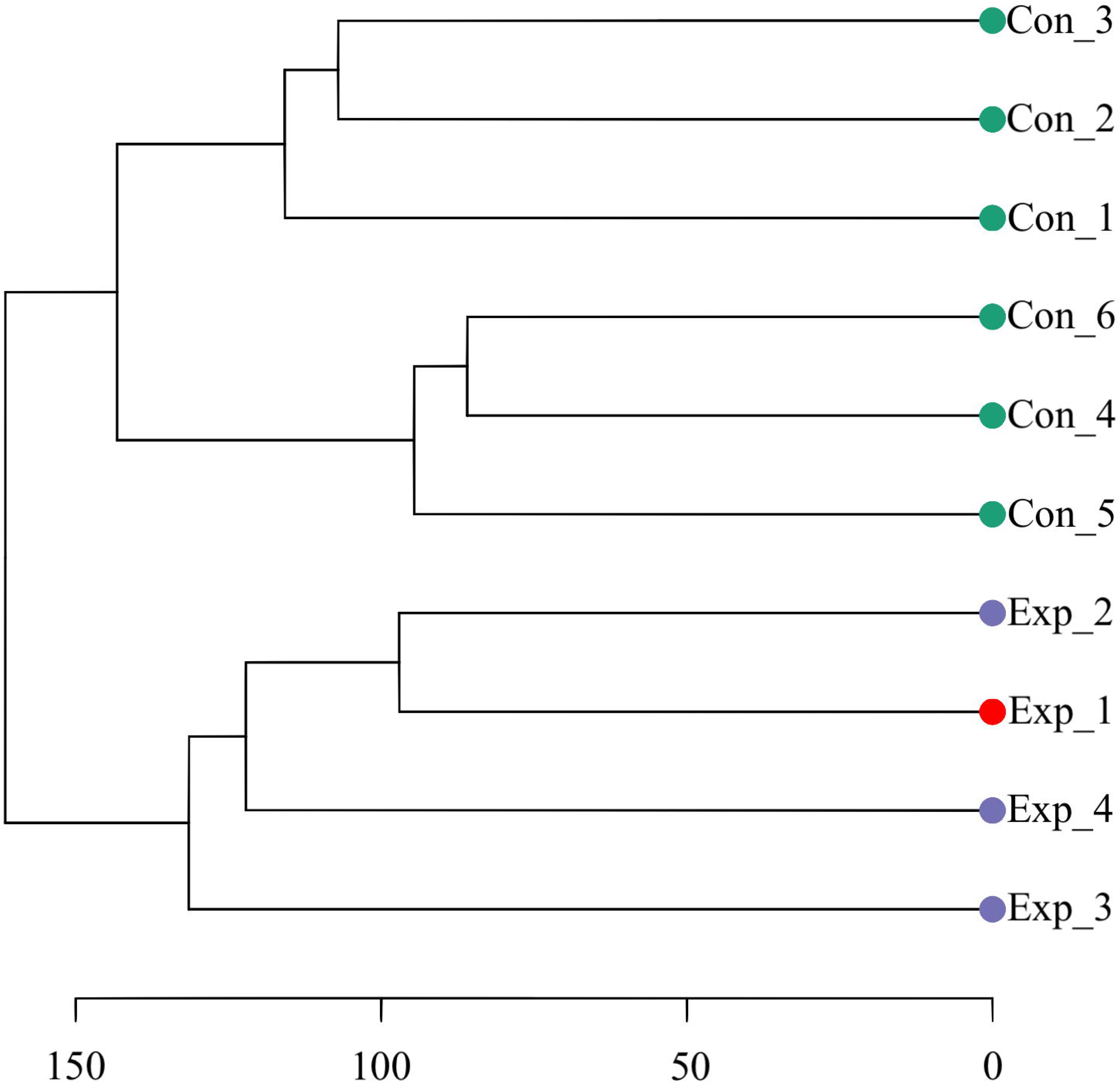
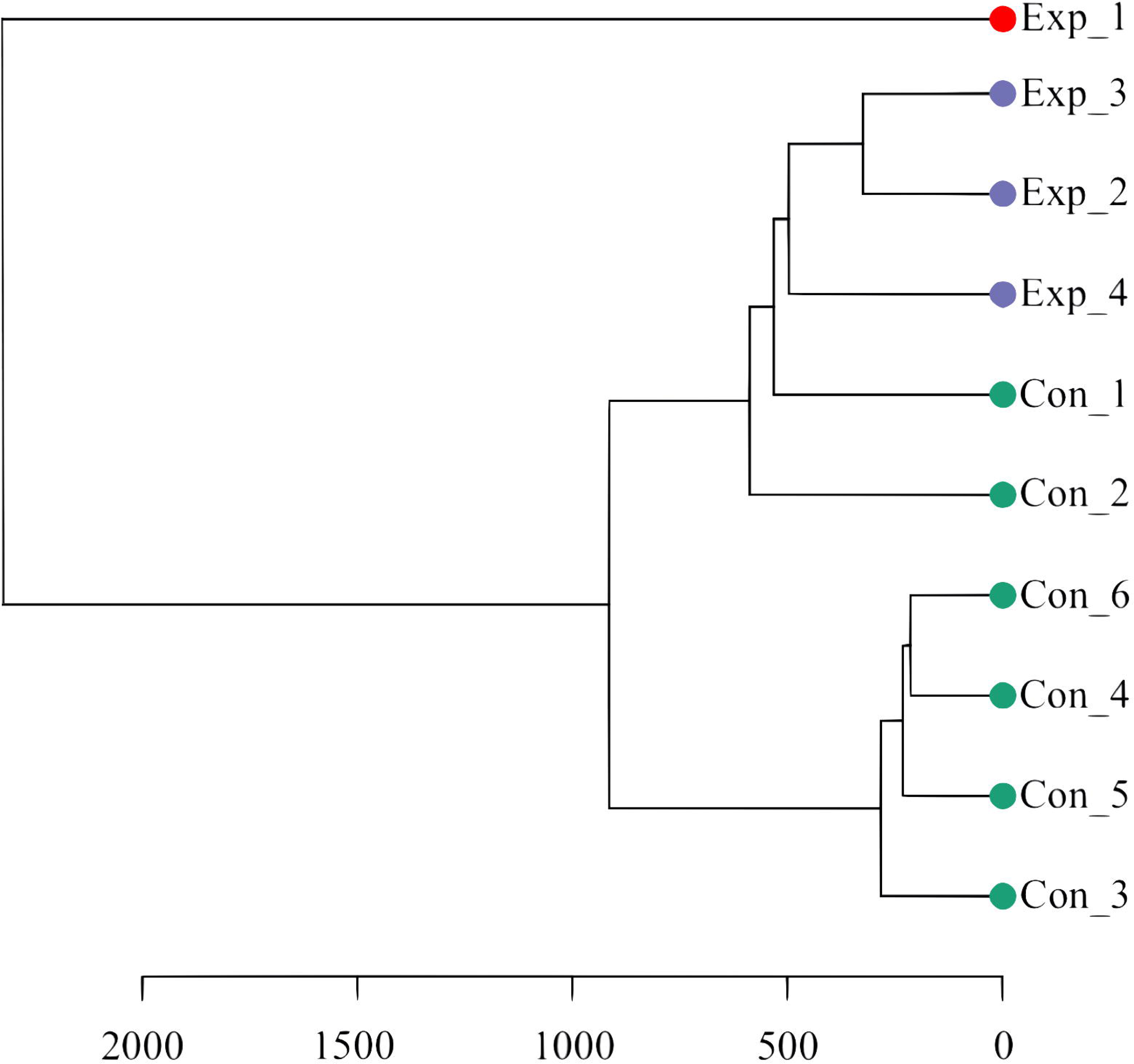
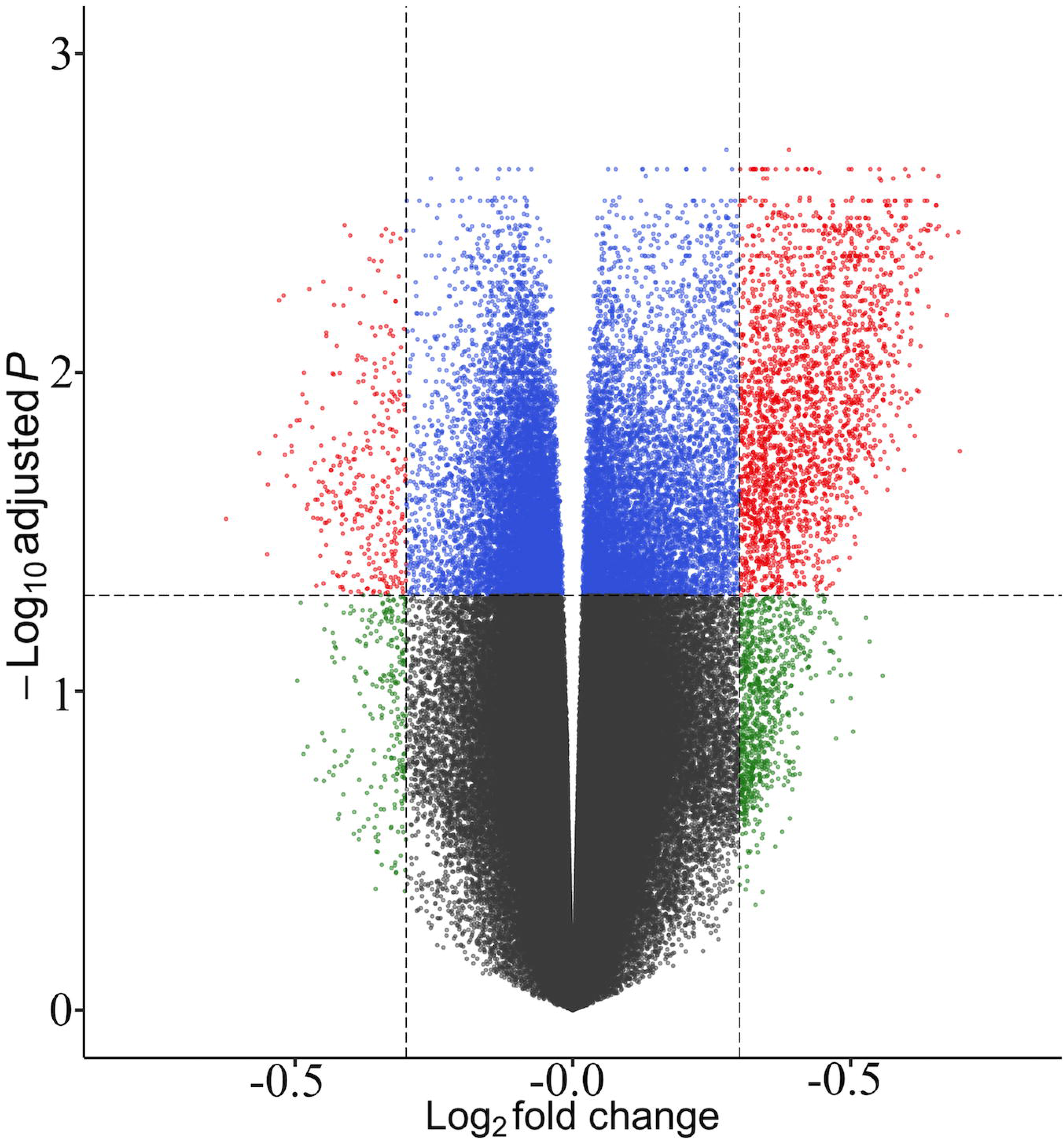
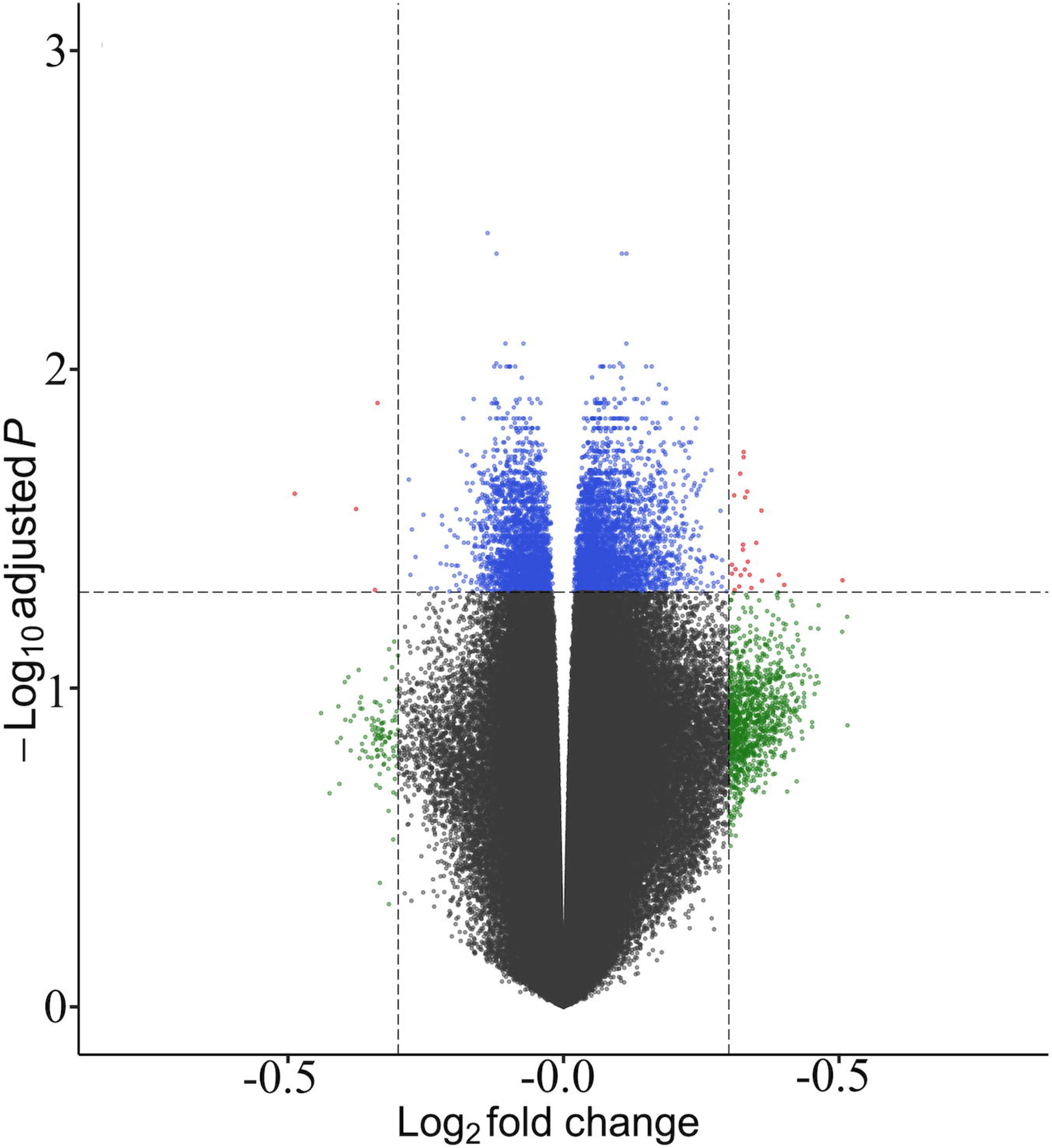
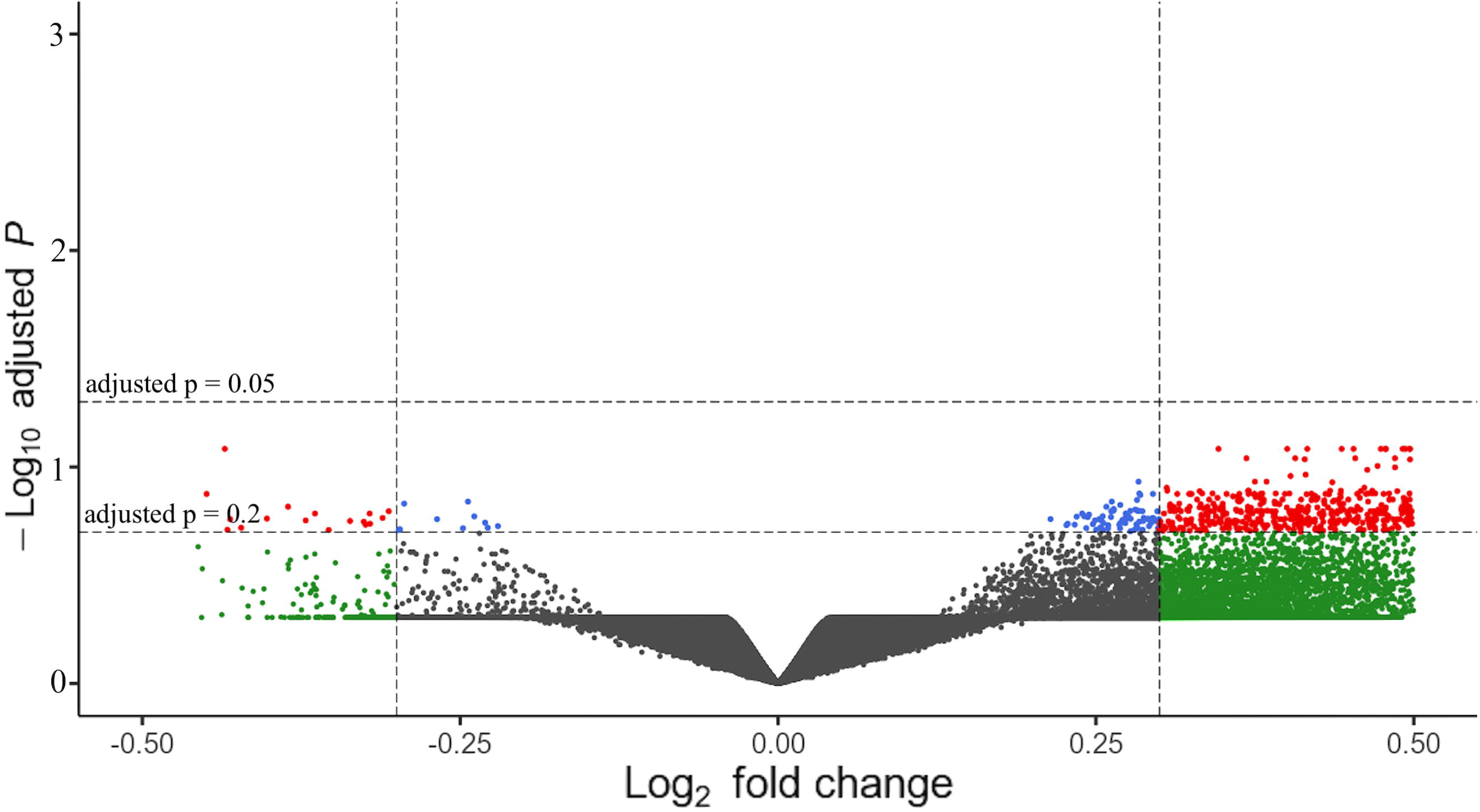
DNA methylome analyses. a, b, c) Dendrograms of the unsupervised hierarchical clustering of the DNA methylation β-values of from HLA-DR, CD3 and PBMCs, respectively. “Con”: green=controls, “Exp”: purple=TB-exposed, red=TB index case. The scale defines the clustering Euclidean distance. d, e, f) Volcano plots of DMGs from HLA-DR, CD3 and PBMCs. Red dots represent DMGs above cut-offs (±0.3 Log_2_ fold change and BH-corrected *p*-value < 0.05, <0.1 or 0.2 as indicated).

Next, we identified the differentially methylated CpG sites (DMCs) and DMGs by comparing the TB-exposed and controls groups for each cell population. To filter out the most significantly altered DMGs in the dataset, the stringency criteria of log_2_ 0.3 fold increased or decreased β-values and Benjamini-Hochberg (BH)-corrected *p*-value < 0.05 (HLA-DR), <0.1 (CD3) and <0.2 (PBMC) were applied. The results are depicted as volcano plots, which show that the DNA methylomes of TB-exposed most strongly differ in the HLA-DR cells as compared to control subjects, followed by the CD3 population, whereas PBMC datasets reveled fewer DMGs (Fig. 2d, e, f, Table 2). To highlight the locus position of the DMGs, chromosome maps were constructed (Suppl. Fig. S1). Using the same stringency criteria as for the HLA-DR analysis, we tested whether DMGs would emerge when the datasets were arranged in other possible groups as derived from the demographics (>/< median age, sex, IGRA status), Neither age nor IGRA status generated any significant DMGs with these settings, and gender rendered only three (Table 2).

**Table 2:**
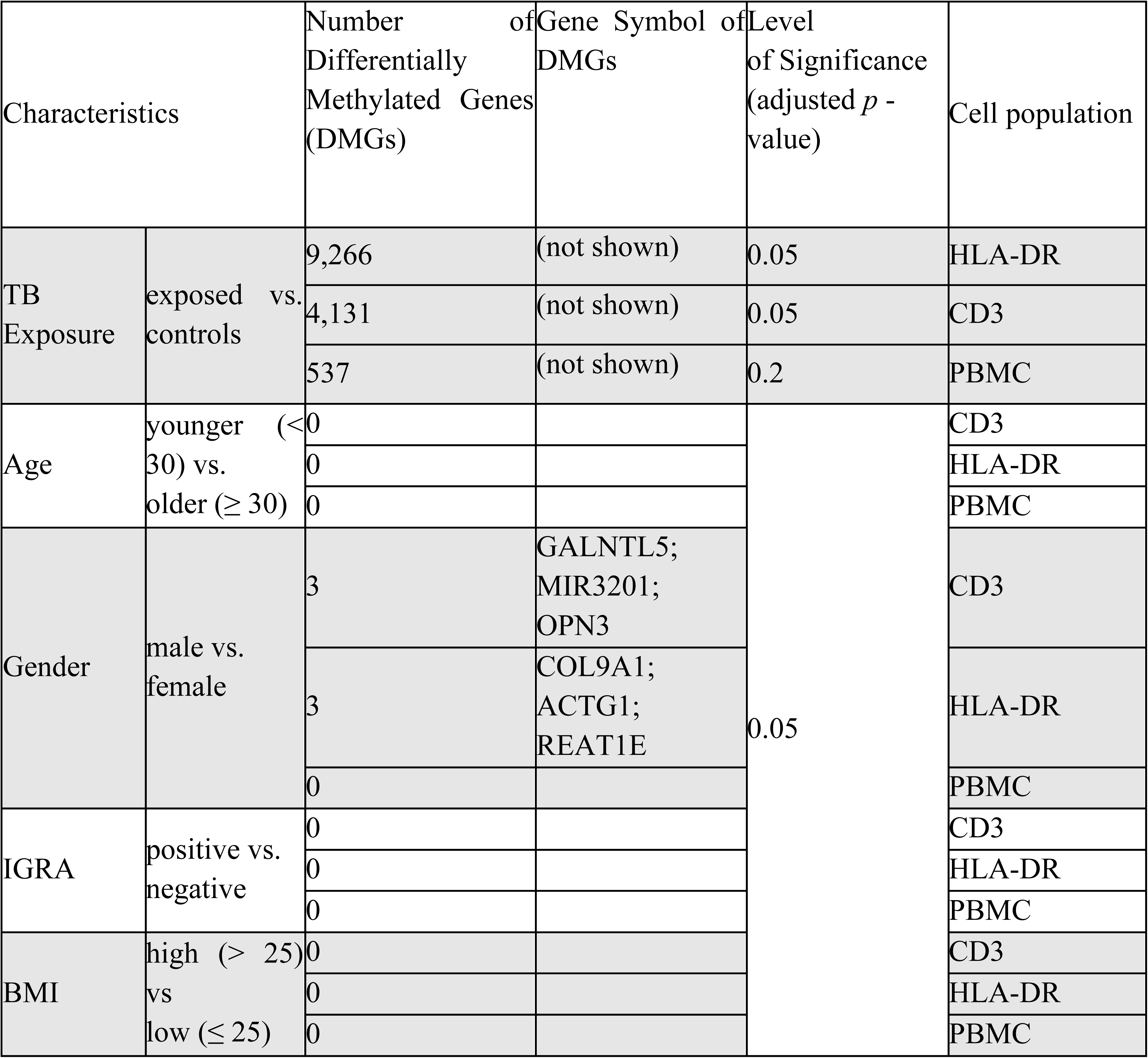
Differentially methylated genes emerging through comparison of different characteristics (TB-exposure, age, gender, IGRA status and BMI) of the participants in the three different cell populations, CD3, HLA-DR and PBMC.

**Table 3:**
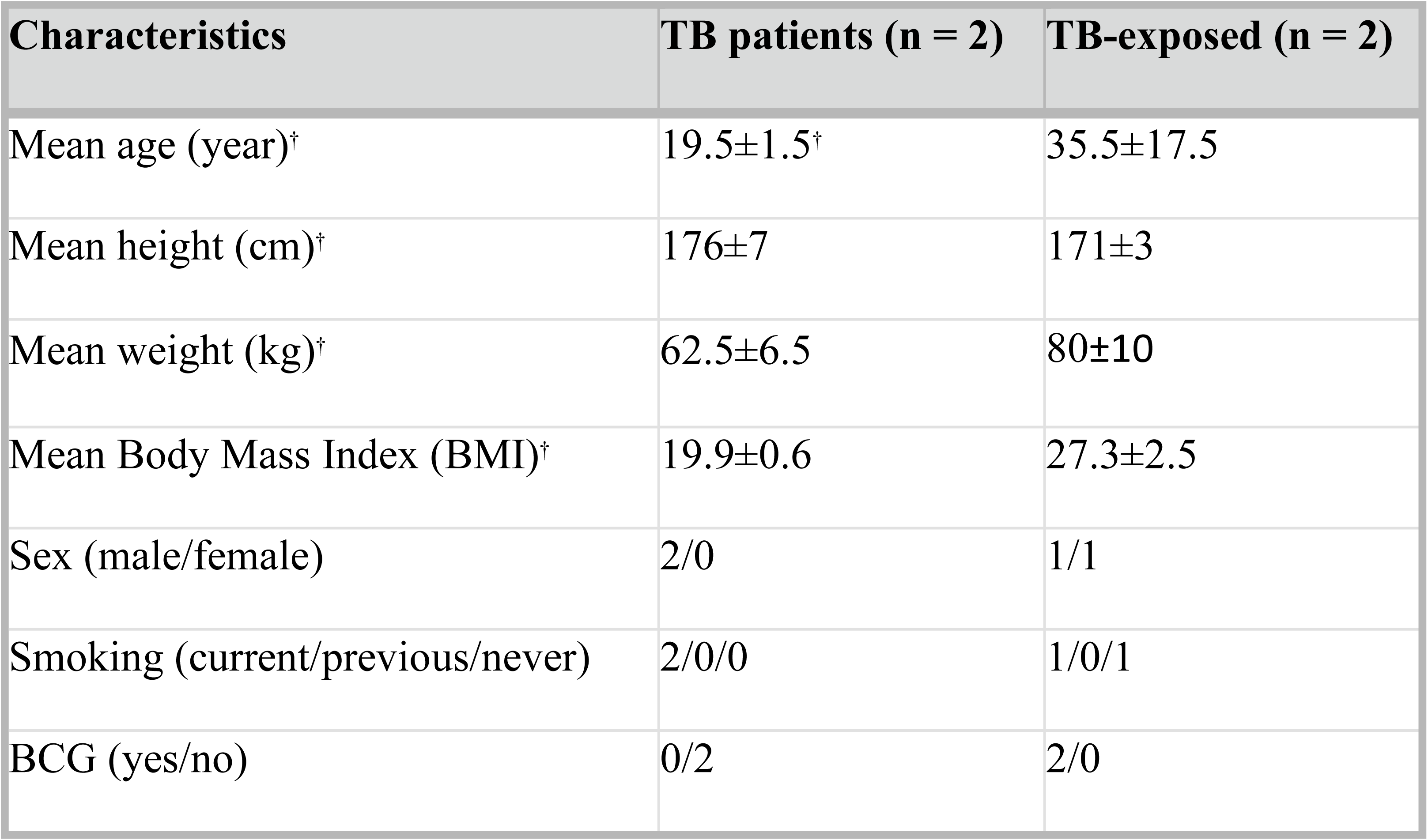
Demographic data of the participants in the second recruitment (“test dataset”)

| Characteristics | TB patients (n = 2) | TB-exposed (n = 2) |
| --- | --- | --- |
| Mean age (year)<sup>†</sup> | 19.5±1.5<sup>†</sup> | 35.5±17.5 |
| Mean height (cm)<sup>†</sup> | 176±7 | 171±3 |
| Mean weight (kg)<sup>†</sup> | 62.5±6.5 | 80±10 |
| Mean Body Mass Index (BMI)<sup>†</sup> | 19.9±0.6 | 27.3±2.5 |
| Sex (male/female) | 2/0 | 1/1 |
| Smoking (current/previous/never) | 2/0/0 | 1/0/1 |
| BCG (yes/no) | 0/2 | 2/0 |

### Functional enrichment analysis reveals common and unique interactomes in the datasets

Using the Panther Database, we investigated whether the identified DMGs were enriched in known pathways (Fig. 3a,b,c). The analysis revealed pathways with relevance for TB infection, including hypoxia-inducible factor (HIF)1-α activation, Vitamin D metabolism and p38, Wnt, Notch, interleukin, chemokine and cytokine signaling pathways ^16–23^. Common pathways shared between at least two of the cell populations included B cell activation, glycolysis, angiotensin II signaling, and cholecystokinin signaling. Notably, several pathways being named after their known functions in the nervous system were enriched in the studied cell populations, including pathways involved in axon guidance and adrenaline, acetylcholine and glutamate signaling. In the PBMC population but not in the lung cell populations, the interferon-γ signaling pathway was identified among the enriched pathways.

**Figure 3.**
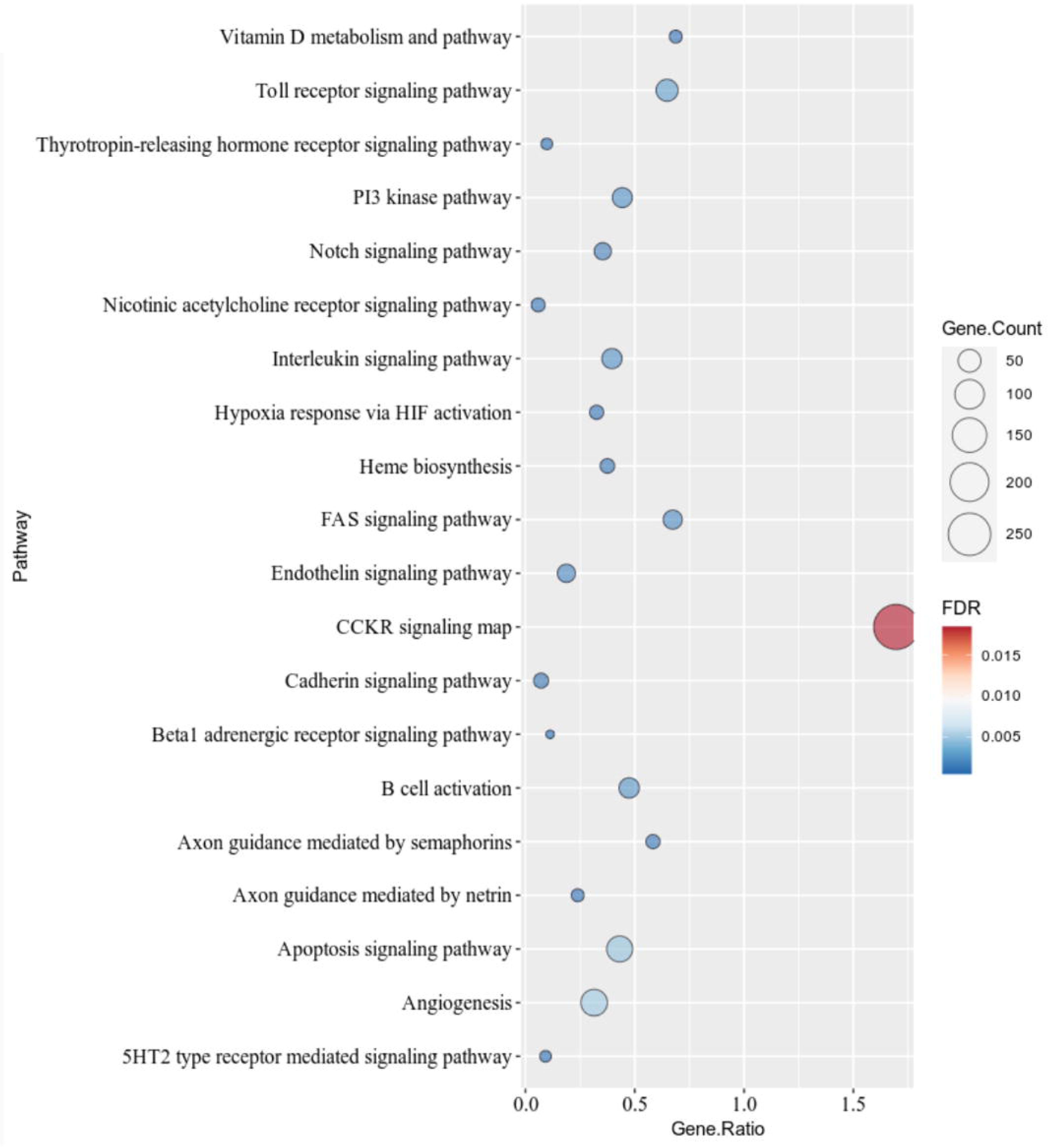
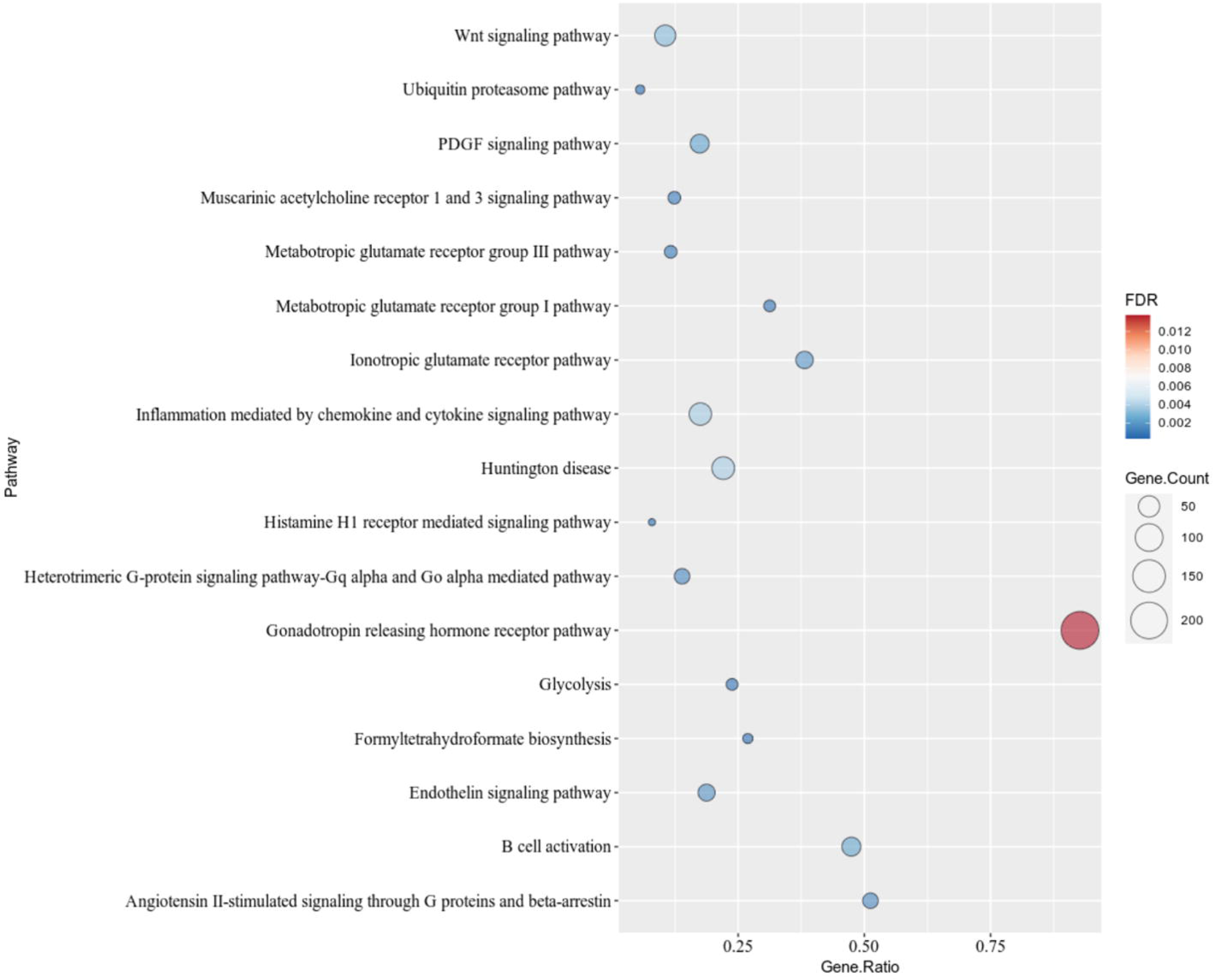
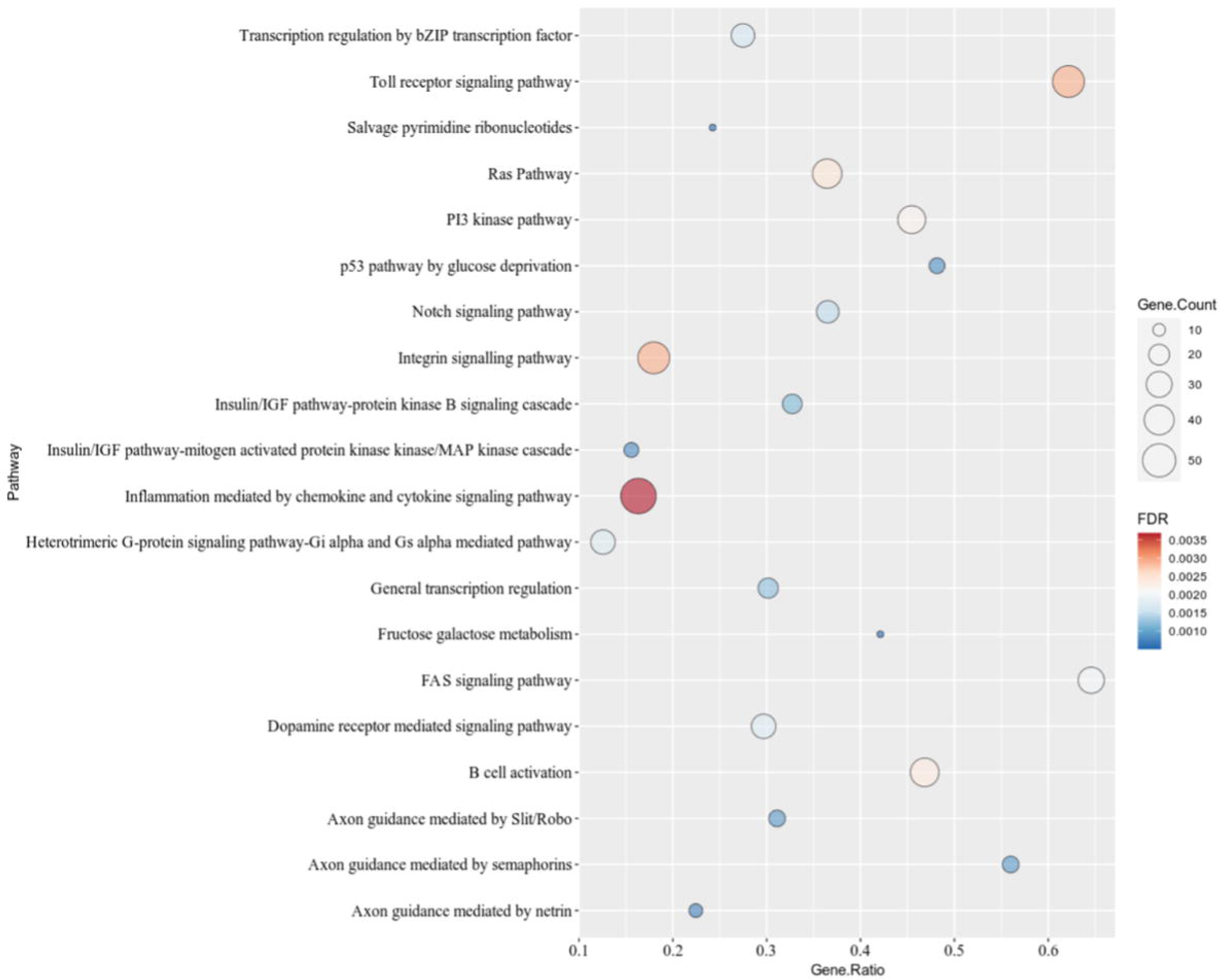
Panther pathway analysis of the identified DMGs with the cut-offs for the different cell populations given in Figure 2. Bubble charts show the gene ratio, gene counts and FDR-corrected *p*-value for a) HLA-DR (top 20 pathways), b) CD3 (totally 17 pathways) and c) PBMC (top 20 pathways).

### Comparisons across cell populations and species reveals the existence of a common DNA methylome-based biosignature in mycobacteria-exposed immune cells

Given the fact that the interaction between mycobacteria and eukaryotes is evolutionary ancient, we predicted that highly conserved pathways exist that are common among the studied cell populations. Combining the identified DMGs from the HLA-DR, CD3 and PBMCs in a Venn analysis, we discovered 185 common DMGs (Fig. 4a). We expanded the Venn analyses to include data from our previous work on BCG vaccine-induced DMGs that correlated with enhanced mycobacterial control^9^, the rationale being that natural exposure to TB and BCG vaccination both represent *in vivo* encounters between mycobacteria and host immune cells. Even though the routes of mycobacterial exposure differ profoundly in these settings, a set of 151 DMGs could be identified that overlapped between our previous BCG study^9^ and all cell populations studied here (Fig. 4b), suggesting that a highly conserved epigenetic response to mycobacterial challenge exists.

**Figure 4:**
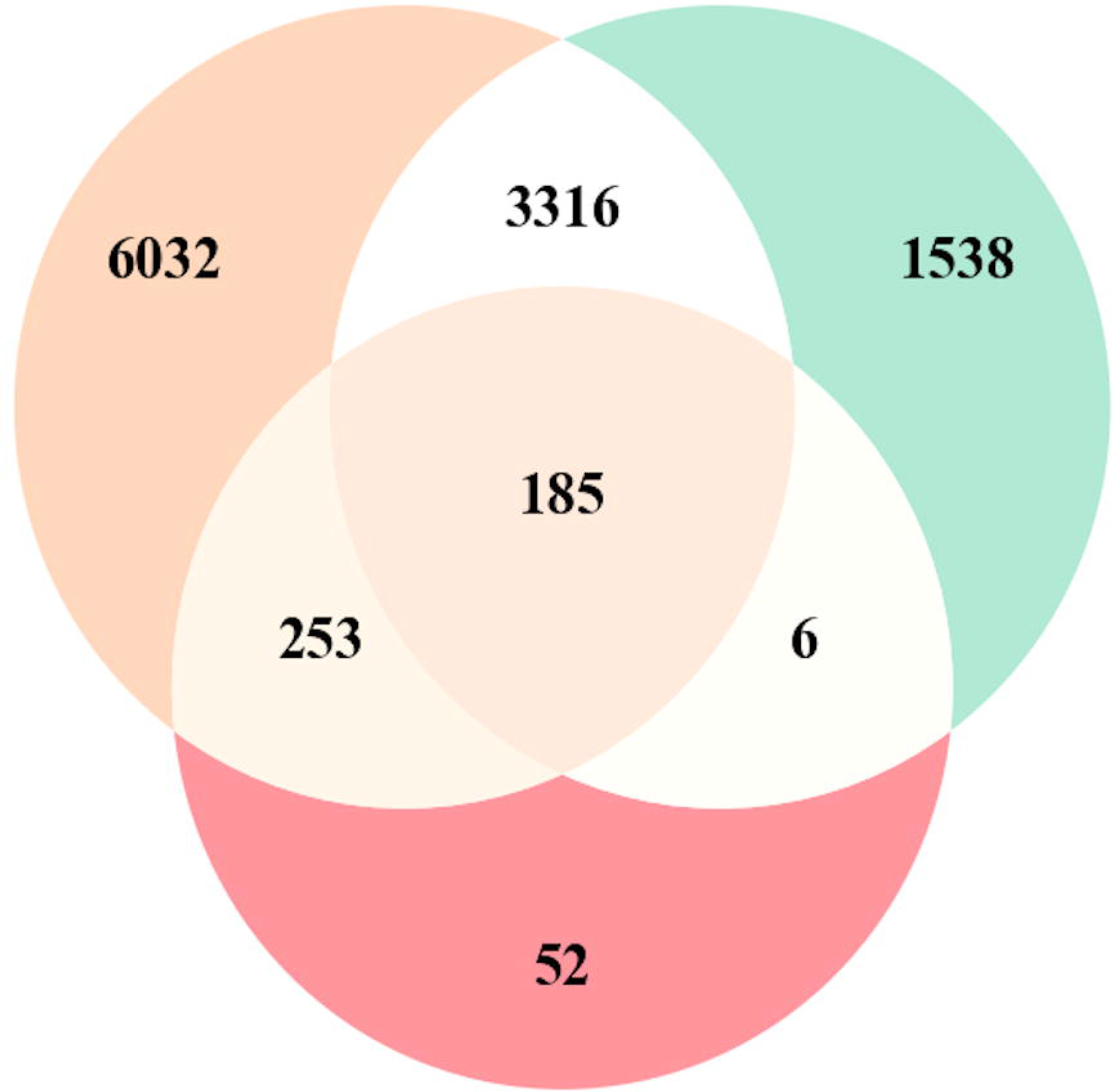
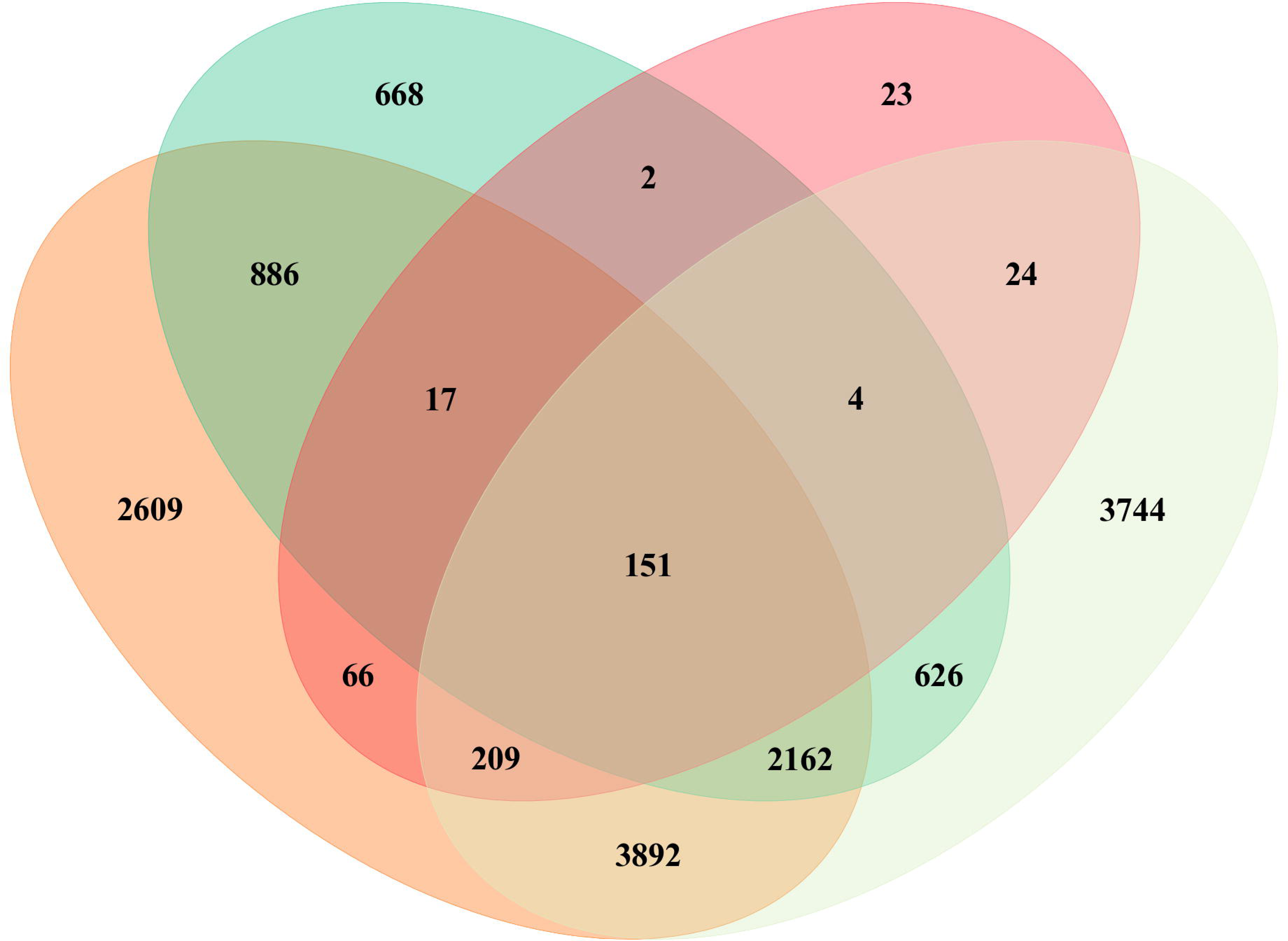
Venn analyses comparing DMGs, pathways and GO terms between different datasets a) Overlapping DMGs derived from the HLA-DR, CD3 and PBMC DNA methylomes, b) Overlapping DMGs from this study and from our previous work (Verma et al).

In 2018, Hasso-Agopsowicz *et al*. described alterations in DNA methylation patterns in PBMCs from BCG-vaccinated individuals, with concomitant enrichment in many immune-related pathways^10^. In order to compare that study with ours, we performed Panther analysis with the 185 common DMGs and matched the identified enriched pathways with those from that study, revealing that 75% of those pathways were the same as in the present study (Fig. 5a and Suppl. Table 1a), further corroborating the relationship of the altered DNA methylation patterns induced through TB exposure and BCG-induced changes. In a recent mouse study by Saeed et al. BCG-induced alterations of the epigenome was correlated to protection against *M. tuberculosis* infection^11^, and to translate our human DNA methylome signature to the signature identified in the mouse study, we searched for pathway overlaps between the two studies. To allow comparison with that study, we performed a Gene Ontology (GO) enrichment analysis (Suppl. Fig. S2a-c). Figure 5b and Suppl. Table 1b demonstrates that for our PBMC data, the GO terms “biological processes” overlapped to 100% with the mouse study (same cell population) and to 31% and 65% for HLA-DR and CD3 cells respectively. In 2014, Saeed et al ^24^ demonstrated the induction of trained immunity pathways by another immune-training agent, β-glucan. We assessed possible pathway overlap with that study and although there were fewer overlaps as compared to the BCG-induced pathways described above, again the strongest correlation was found in the PBMC fraction, in this case in the GO terms “cellular components” (Fig. 5c and Table 1c).

**Figure 5.**
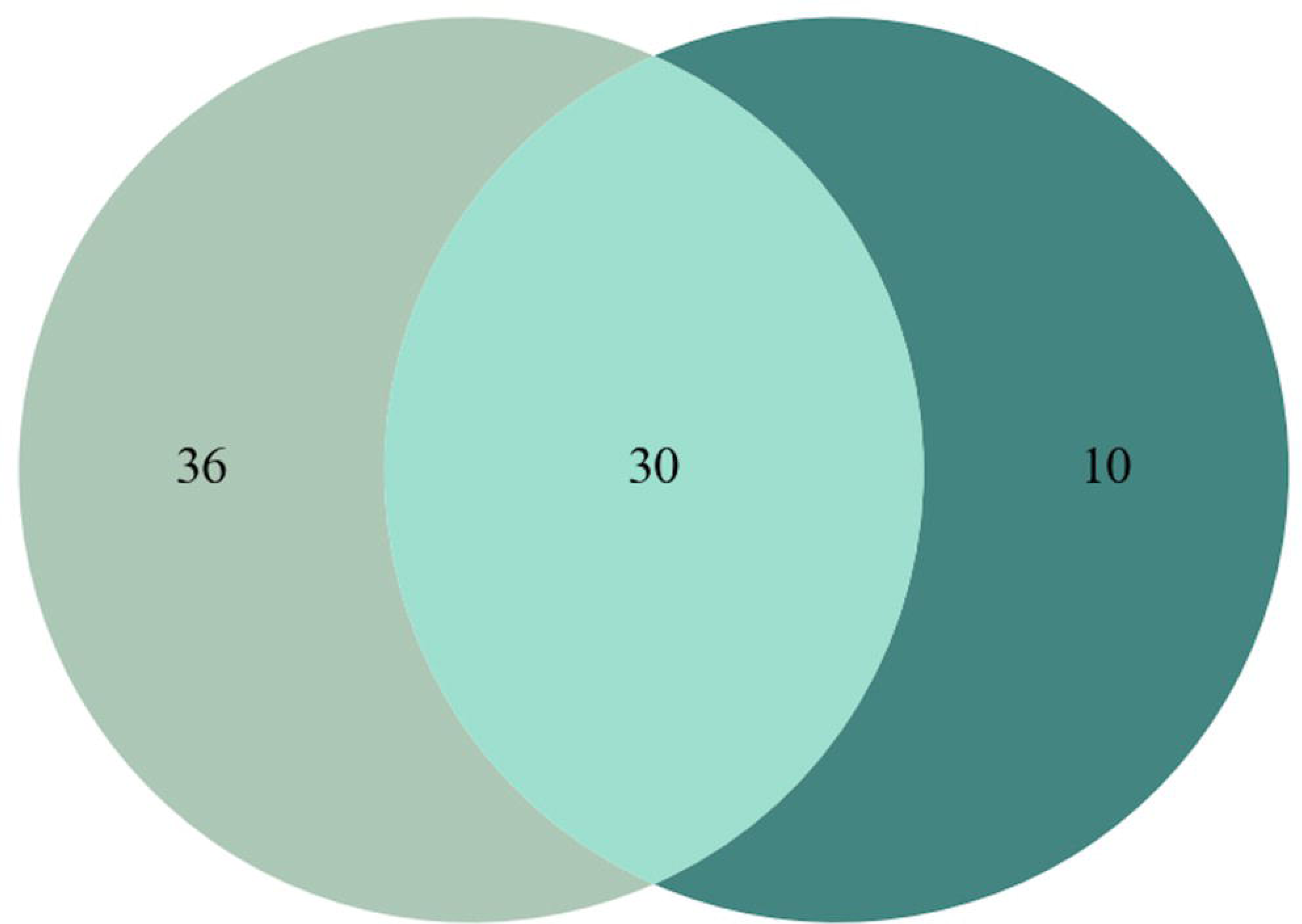
Pathway overlap with other studies’ results. a) Venn diagram describing the number of Panther pathways overlapping between the ones derived from the 185 common DMGs in this study (dark green) and Hasso-Agopsowicz et al (human BCG vaccine study, light green) ^10^, b) Sunburst Plot describing the overlap of enriched GO biological processes emerging from a comparison between the GO data derived from the 185 common DMGs (Figure 4a) and Kaufmann et al (BCG study performed in mouse PBMCs^11^, c) Sunburst Plot describing the overlap of enriched GO biological processes emerging from a comparison between the GO data derived from the 185 common DMGs and Saeed et al (study on trained immunity induced by β-glucan^24^).

Finally, we assessed how well the 284 CpG sites corresponding to the 185 overlapping DMGs performed in an unsupervised cluster analysis. To this end, we included one additional TB patient and two contacts, and collected HLA-DR cells from induced sputum, since the DNA methylome data this cell type was clearly outperforming the others with respect to accurate separation of the groups. Fig. 6 shows a *k* means-based dendrogram with a heatmap of the β values of the 284 CpG sites from the previous and the new subjects’ samples.

**Figure 6.**
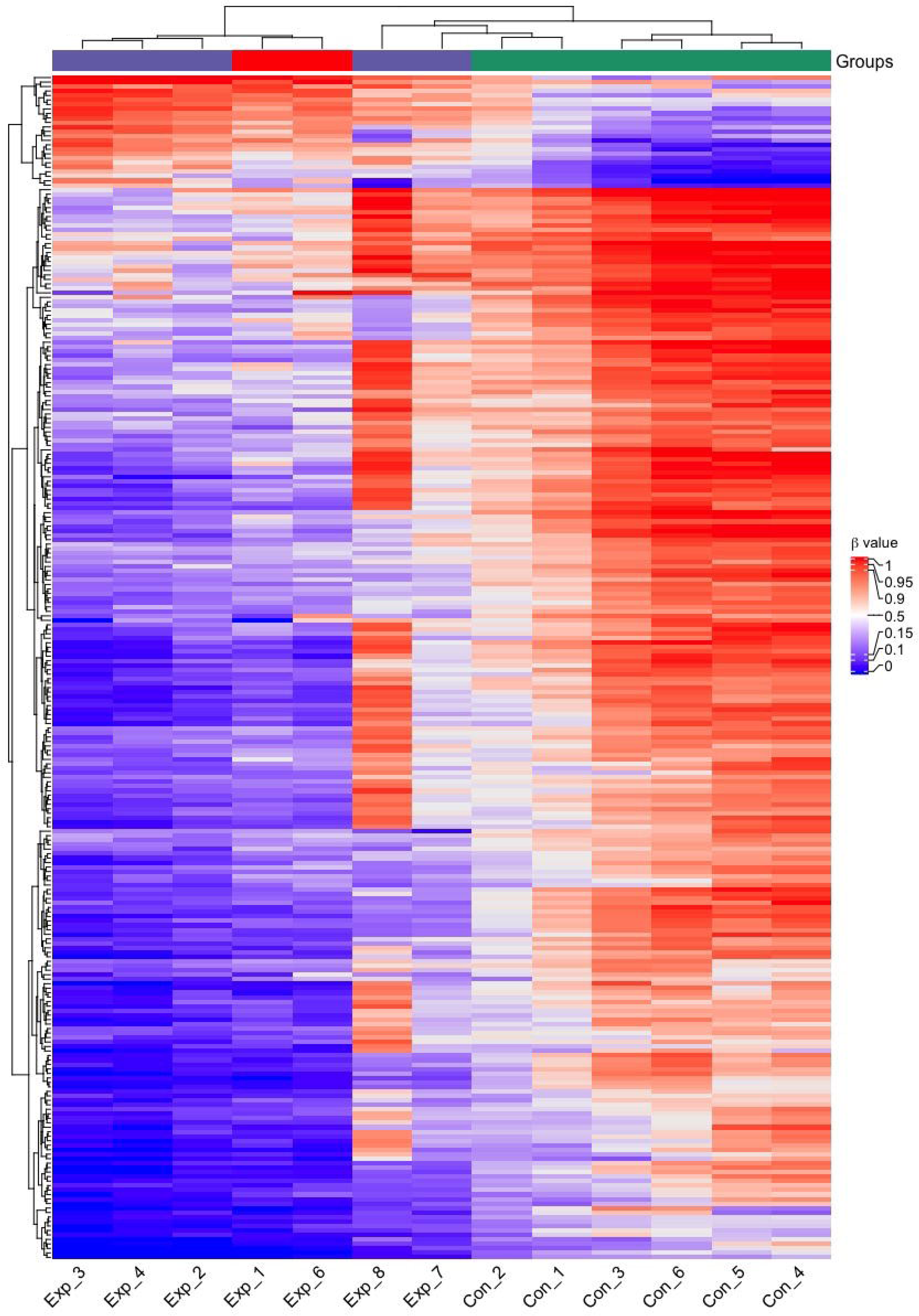
Heatmap of the HLA-DR-derived β values of the signature’s 284 CpG sites of the 6 initial subjects (Exp_1-4 and Con_1-6) and the three additional exposed subjects (Exp_6-8). Purple=exposed, red=TB index case, green=controls.

Together the results demonstrate that the identified biosignature with the strongly enriched pathways can be linked to modifications of immune cell functions that result in improved anti-mycobacterial defense, possibly through trained immunity.

## Discussion

In this study, we present data suggesting that exposure to TB generates a very distinct DNA methylation signature in pulmonary immune cells. The signature was found not only in those with active or latent TB infection but also in individuals who are exposed but IGRA-negative. The finding that also healthy, TB-exposed carry the signature opens up the possibility that the epigenetic alterations reflect a host-beneficial reprogramming of the immune mechanisms rather than being induced by *M. tuberculosis* as a step to evade the immune defense. This notion is supported by our observation that the DMGs identified in the present study strongly overlapped with the previously reported DNA methylation changes induced during BCG vaccination, which correlated with increased anti-mycobacterial capacity of macrophages^9^. In addition, we demonstrate that the GO data derived from our dataset display a strong overlap with data from a study on protective BCG vaccination in mice^11^.

BCG vaccination has convincingly been shown to induce heterologous immunity protecting against childhood mortality from other causes than TB^25, 26^. Based on our finding that natural TB exposure and BCG vaccination trigger similar epigenetic changes we propose the hypothesis that a “beneficial exposure” to TB exists, which protects against other infections through heterologous immunity. Along the same line, it has been shown that a substantial fraction of individuals exposed to TB can be defined as ‘early clearers’, since they do not test positively in the tuberculin skin test (TST) or IGRA^27^, suggesting effective eradication of the infection ^27^. Identifying these early clearers and understanding the biology behind their resistance to TB infection could move the field forward towards novel strategies of TB prevention.

The rationale of comparing overlaps in mycobacteria-induced DNA methylation changes between different immune cells is based on the accumulated evidence of co-evolution of mycobacteria and amoeba ^28, 29^ and the origin of phagocytic immune defense in metazoans from amoeboid cells^30, 31^, predicting evolutionary conserved pathways to be engaged in anti-mycobacterial defense. In line with this prediction, we identified the ubiquitously expressed and evolutionary conserved Wnt signaling pathway, which is found in all metazoans^32^ and with homologs in amoeba^33^ to be strongly enriched across all cell populations, settings and species. The role for Wnt signaling in mycobacterial defense remains elusive, but many studies have ascribed Wnt pathways immunomodulating functions and induction during *M. tuberculosis* infection (reviewed in ^20^).

In concordance with the macrophage being the main host cell for mycobacterial infection, the strongest enrichment of DNA methylation changes was observed in the HLA-DR-positive cell population, which is dominated by alveolar macrophages. The pathways identified to be enriched in the HLA-DR population have been described in the context of trained immunity, BCG exposure and TB. For example, activation of Hypoxia-Inducible Factor 1 α and glycolysis pathways (P00030 and P00024, respectively) are hallmarks of macrophages that have undergone the epigenetic changes that are reflective of trained immunity (reviewed in ^34, 35^), which is induced in myeloid cells upon BCG-stimulation^36^. VEGF-release (P00056) by macrophages has been shown to recruit immune cells during granuloma formation ^37^. Further, vitamin D has been shown to strengthen the anti-mycobacterial activity of macrophages^17, 38^, and upregulation of components of the vitamin D pathway is linked to the production of anti-microbial peptides^18^, providing a possible effector mechanism for mycobacterial control. Recent literature on immune regulation through T cell-derived acetylcholine^39, 40^ attributes relevance to the acetylcholine receptor pathway identified among the HLA-DR pathways.

Although macrophages and lymphocytes are not generally viewed as having many similarities, we found 34 of the identified pathways to overlap between HLA-DR and CD3. In data derived from the CD3 and PBMC populations, both of which represent lymphocytes, overlaps were identified for glycolysis, glutamate receptor and angiotensin II pathways. Interestingly, a metabolic shift towards increased glycolysis, representative of the Warburg effect, has been strongly associated with trained immunity ^34^. However, the literature is dominated by the view that this event takes place in trained myeloid cells, while we identified this circuit in CD3 cells (lymphocytes) and not in the HLA-DR cells (dominated by macrophages). The glutamate receptor is widely expressed on immune cells and have been described as having an important regulatory role in T cells, which can also produce and release glutamate^41^. The role for angiotensin II pathway in TB remains elusive, while Angiotensin II Converting Enzyme 2 is currently in the spotlight due to fact that the SARS-CoV2 virus utilizes it as a receptor for entry into host cells^42^. In the PBMC population, which over all showed a weaker epigenetic response, we found the interferon-γ signaling pathway, which has a well-established role in anti-mycobacterial defense (reviewed in ^43^), to be among the reprogramed pathways.

A weakness of our study is the small cohort, which warrants testing in larger cohorts performed in different clinical settings such as areas high- and low endemic for TB. Taken together, we present data supportive of DNA methylation changes that are induced through exposure to TB. The changes correlate with findings from studies on BCG vaccination including TB protection, heterologous and trained immunity.

## Methods

### Study design and participants

Patients with pulmonary TB, participants with occupational- or household-related TB-exposure and healthy controls, with an age ranging from 18 to 53 years, were enrolled at Linköping University Hospital and Linköping University, respectively. Included subjects (please see Table 1 for demographics) donated peripheral blood and induced sputum samples^12^ following oral and written informed consent (ethical approval obtained from the regional ethical review board in Linköping, #2016/237-31). The study protocol included questionnaires on respiratory and overall health, the evaluation of IGRA-status and sputum samples for DNA extraction.

### Induced sputum and pulmonary immune cell isolation

Induced sputum is a well-tolerated, non-invasive method to collect cells from the surface of the bronchial airways after inhalation of a hypertonic saline solution. The procedure of sputum induction takes approximately 30 minutes and is both cost effective and safe with minimal clinical risks^44^. Sputum specimens were collected as described by Alexis et al^45^, with the following modifications: premedication with an adrenergic β2-agonist, salbutamol (Ventoline, 1ml 1mg/ml) was administrated before the inhalation of hypertonic saline, using a nebulizer (eFlow, PARI). The subsequent steps of sputum processing were adopted from Alexis et al. (2005)^46^ and Sikkeland et al. ^14^. The HLA-DR and CD3-positive cells were isolated using superparamagnetic beads coupled with anti-human CD3 and Pan Mouse IgG antibodies and HLA-DR/human MHC class II antibodies (Invitrogen Dynabeads, ThermoFisher, cat no 11041 and 14-9956-82, respectively). An initial positive selection was done with CD3 beads followed by a positive HLA-DR selection. Bead-coating and cell isolation was performed according to manufacturer’s protocol.

### PBMC isolation

Following venipuncture, PBMCs were isolated using SepMate −50 tubes (StemCell Technologies), as per the manufacturer’s protocol. The blood from each subject was poured into 50 ml tubes (Falcon, ThermoFisher) and carefully diluted 1:1 with a Dulbecco’s PBS (D-PBS, Gibco by Life Technologies) containing 2% fetal bovine serum solution. Then, 15 ml Lymphoprep (StemCell Technologies) was transferred into a SepMate −50 tube (StemCell Technologies) and the diluted sample subsequently added to the Lymphoprep-filled tube, by carefully pipetting down the side of the tube. After centrifugation (1 200 x g, 10 minutes, RT), the plasma layer was removed and the PBMCs poured off into a sterile 50 ml tube (Falcon, ThermoFisher) containing D-PBS (Gibco by Life Technologies). The tube was subsequently filled with cold D-PBS until a total volume of 50 ml and centrifuged (300 x g, 10 minutes, 4°C). The supernatant was removed, and the pellet gently resuspended in D-PBS (Gibco by Life Technologies), using a transfer pipette (Sarstedt). The sample was then filled up to 50 ml with cold D-PBS, centrifuged (220 x g, 5 minutes, 4°C), and the supernatant discarded. In parallel, IGRA status was determined on whole-blood samples using QuantiFERON-TB Gold (Cellestis) following the manufacturer’s instructions.

### DNA methylation data analysis

DNA of the HLA-DR, CD3 and PBMC was extracted using the AllPrep DNA/RNA Mini Kit (Qiagen) according to the manufacturer’s instructions. Genome-wide DNA methylation analysis was performed using the HumanMethylation450K BeadChip (Illumina, USA) array at the Bioinformatics and Expression Analysis Core Facility at Karolinska Institute, Stockholm. The methylation profiles for each cell type were analyzed from the raw IDAT files in R (v4.0.2) using the *minfi* (v1.36.0) with subset-quantile within array (SWAN) normalization ^47, 48^ and *ChAMP* (v2.19.3) with beta-mixture quantile normalization (BMIQ) ^49, 50^ packages. The type 1 and type 2 probes were normalized using the quantile normalization method. Using the default setup of the *ChAMP* package, following probes were filtered out: i) probes below the detection p-value (>0.01), ii) non-CpG probes, iii) multi-hit probes, and iv) all probes of X and Y chromosomes. Cell type heterogeneity was corrected for the PBMC cell types using the Houseman algorithm ^51^ and batch effects were fixed using *ComBat* from the *SVA* package (v3.38.0) ^52^ Differential methylation analysis were performed with the linear modeling (lmFit) using the *limma* package (v3.46.0) ^53^ in a contrast matrix of the TB-exposed and TB-non-exposed (Control) individuals. All Differentially methylated CpGs (DMCs) were considered significant at the Bonferroni-Hochberg (BH) corrected *p*-value < 0.05 (for HLA-DR cell types), <0.1 (for CD3 cell types) and <0.2 (for PBMC cell types).

### Unsupervised cluster analysis

Hierarchical clustering of the all TB-exposed and control individuals was performed with the normalized β-values obtained after the data filtration in each cell type individually. The distance was calculated using the Euclidean distance matrix. The *dendextend* (v1.14.0) ^54^ and *ape* (v5.4-1)^55^ packages in R were used to construct the horizontal hierarchical plots from the three different cell populations using the *hclust* and *dendrogram* functions.

### Structural annotations

The *EnhancedVolcano*^56^ package (v1.8.0) was used to generate the individual volcano plots from all cell populations. The *ChromoMap*^57^ package (v0.3) was used to annotate and visualize the genome-wide chromosomal distribution of the DMGs. The interactive plots were generated using the *plotly* (v4.9.3) package ^58^.

The heatmaps were generated from the filtered DMGs with their respective CpGs for each cell type using the *ComplexHeatmap* (v2.6.2) package. The clustering dendrogram in heatmaps were plotted using the Euclidean distance matrix.

### Pathway and functional enrichment analyses

We also used Panther database (PantherDB v15)^59^ to identify the enriched pathways related to our identified DMGs. In addition, to assess functional enrichment, we used the *ReactomePA* (v1.34.0)^60^ package with 1000 permutations and the BH-corrected *p*-values. Within the package, GO and Kyoto Encyclopedia of Genes and Genomes (KEGG) were used and using *clusterProfiler* (v3.18.1)^61^, we performed KEGG pathway enrichment analysis (data not shown). To enhance the visualization and better understanding of the enrichment result, *GOplot* (v1.0.2)^62^, another R package was used. The pathway enrichment were also calculated using the topology-based ontology methods using *RontoTools*^63^ (v2.18.0)^64^ (v2.40.1)*, SPIA*^65^ (v2.42.0) and *pathview*^66^ (v1.30.1) was used to visualize the related pathways with the KEGG pathway maps (data not shown).

### Venn and overlap analyses

Venn analyses were performed in order to detect the DMGs overlapping between cell populations and between studies. We constructed the Venn diagrams by using *matplotlib-venn* package (https://github.com/konstantint/matplotlib-venn) using in-house python script. The overlap analyses were calculated and plotted using the *go.Sunburst* function from plotly using an in-house python script.

### Statistical analyses

All differences with a *p-*value < 0.05 were considered significant if not otherwise stated. We calculated family-wise error rate (FWER) using the BH correction method. All analyses were performed in R (v4.0.2) with the mentioned packages.

## Data Availability

Sequencing datasets will be made publicly available upon acceptance and prior to final publication.

## Acknowledgements

This study was funded through generous grants from Forskningsrådet Sydöstra Sverige (FORSS-932096), the Swedish Research Council (2015-02593 and 2018-02961) and the Swedish Heart Lung Foundation (20150709 and 20180613). J.D is a postdoctoral fellow supported through the Medical Infection and Inflammation Center (MIIC) at Linköping University. We direct our gratitude to the staff at Linköping University Hospital and the Vrinnevi Hospital in Norrköping for assistance in sample collection and all the subjects for donating samples. The DNA methylome data were generated at the Bioinformatics and Expression Analysis Core Facility at the Department of Biosciences and Nutrition, which is supported by the Board of Research at the Karolinska Institute, Stockholm. The computations were enabled by resources provided by the Swedish National Infrastructure for Computing (SNIC) at Linköping University campus partially funded by the Swedish Research Council through grant agreement no. 2018-05973.

## Author contributions

M.L., N.I. and J.P. conceived and designed the study, N.I. and I.P. collected the patient samples and compiled the demographics tables, J.D. and M.L. designed and performed the bioinformatic analysis of the data, J.D. wrote the code and created the figures. N.I, I.P. J.D and M.L wrote the manuscript. All authors declare no conflict of interest.

---

**Supplementary Figure S1.**
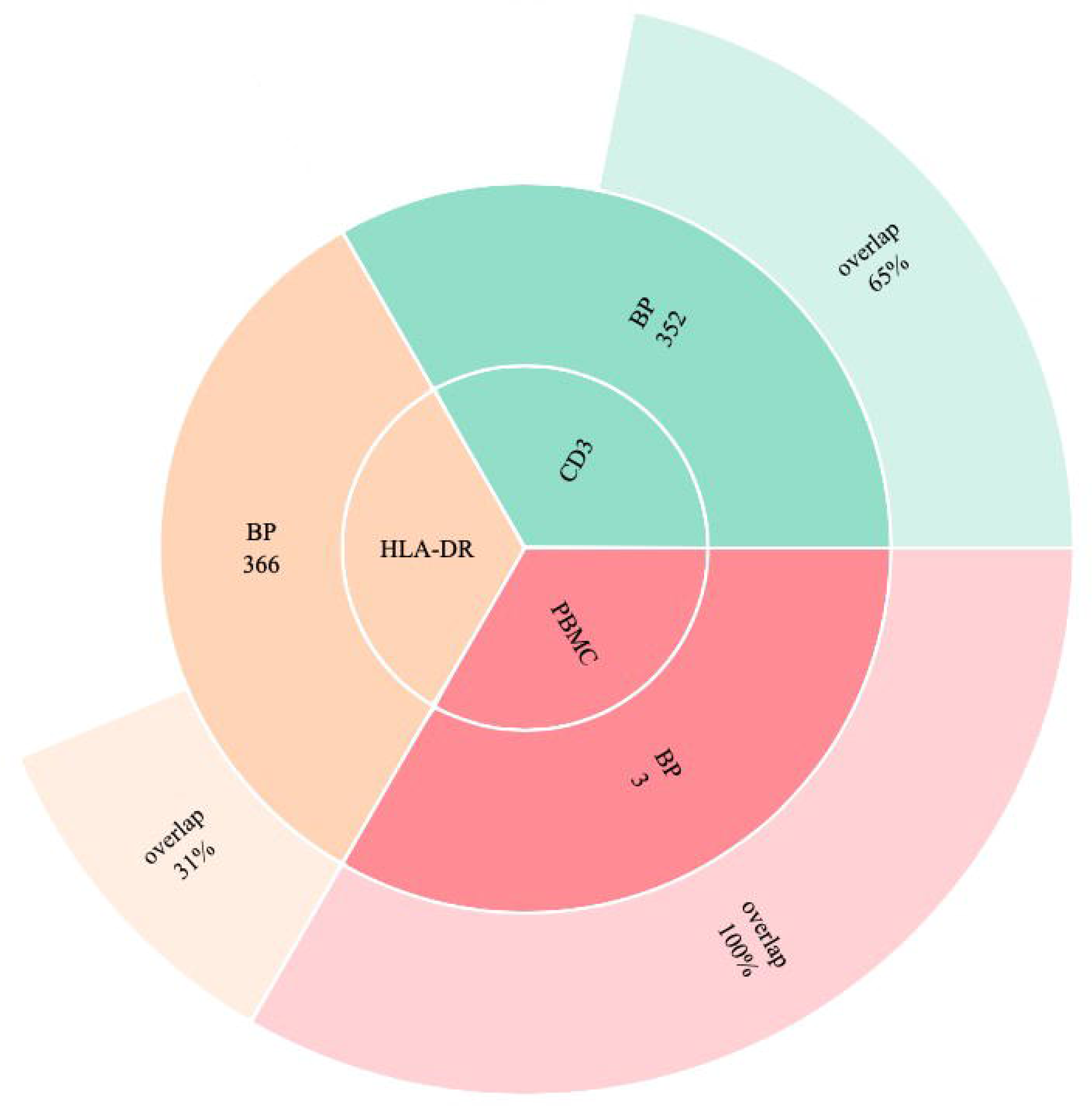
Chromosome maps showing the chromosomal distribution of all DMCs in 22 autosomes (hypomethylated regions in blue and hypermethylated regions in red). a) HLA-DR, b) CD3 and c) PBMC.

**Supplementary Figure S2.**
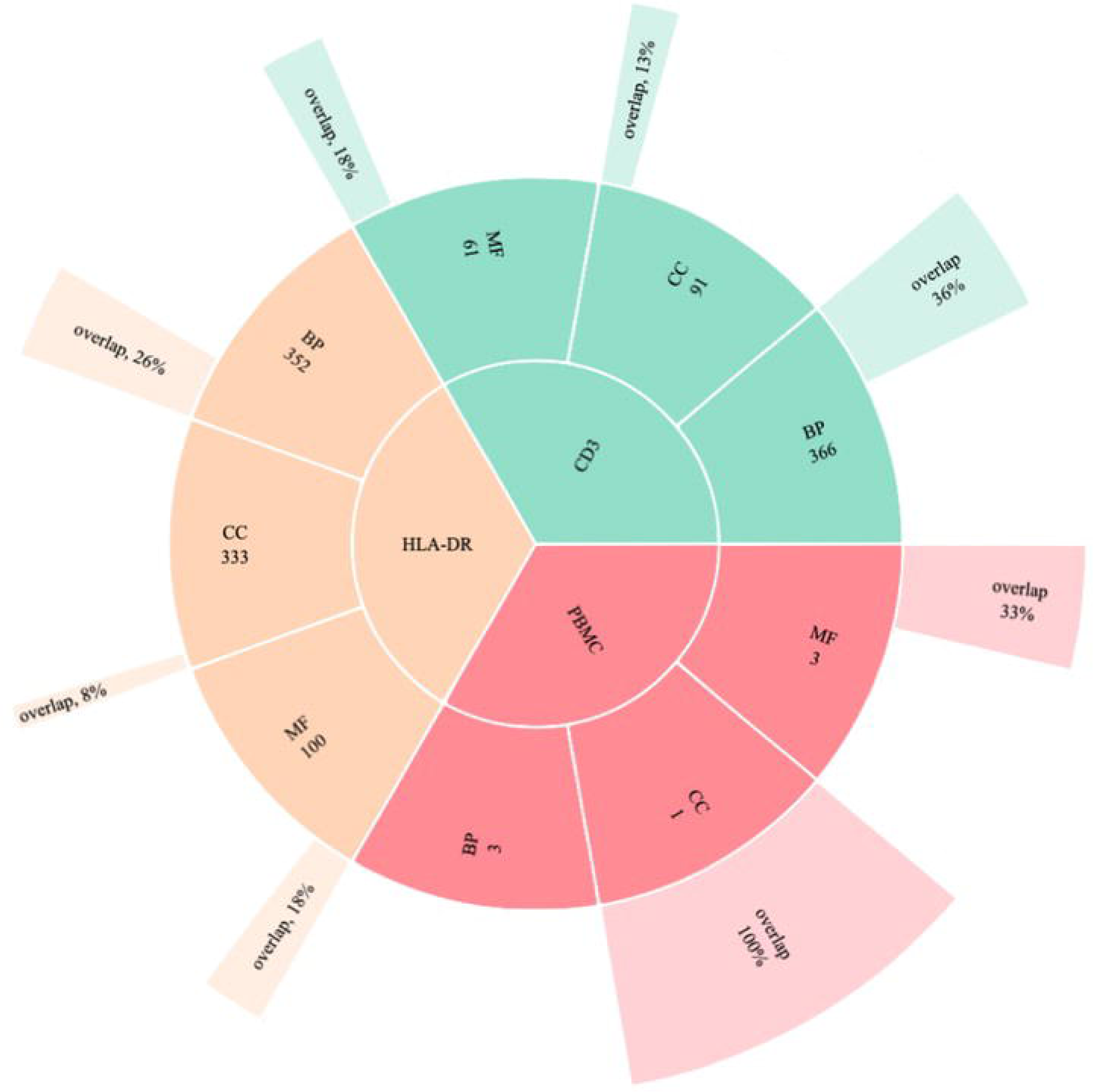
Gene Ontology (GO) enrichment functional analysis a) HLA-DR, b) CD3, c) PBMC and d) the list of functional categories identified in the dataset. The colour bar shows the logFC values ranging from −1 (blue) to 1 (red).

