## Supplementary figures and images for "A DNA methylome biosignature in alveolar macrophages from TB-exposed individuals predicts exposure to mycobacteria"

### S1a

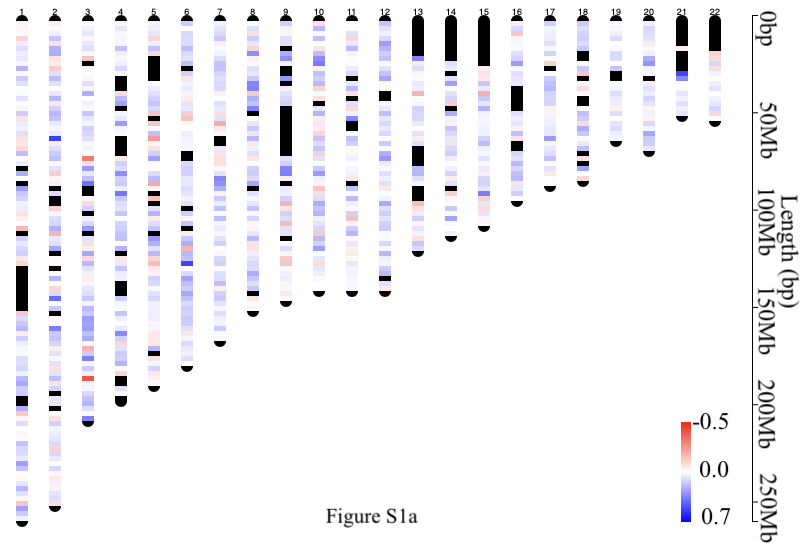

### S1b

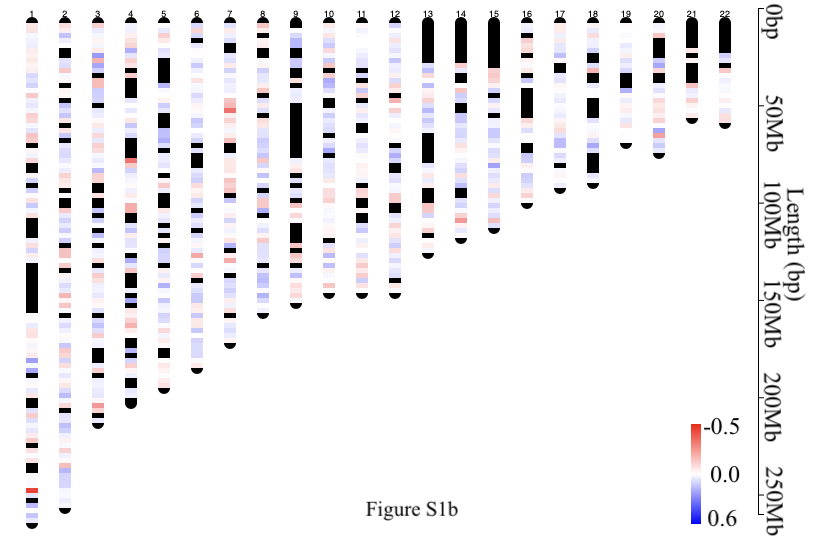

### S1c

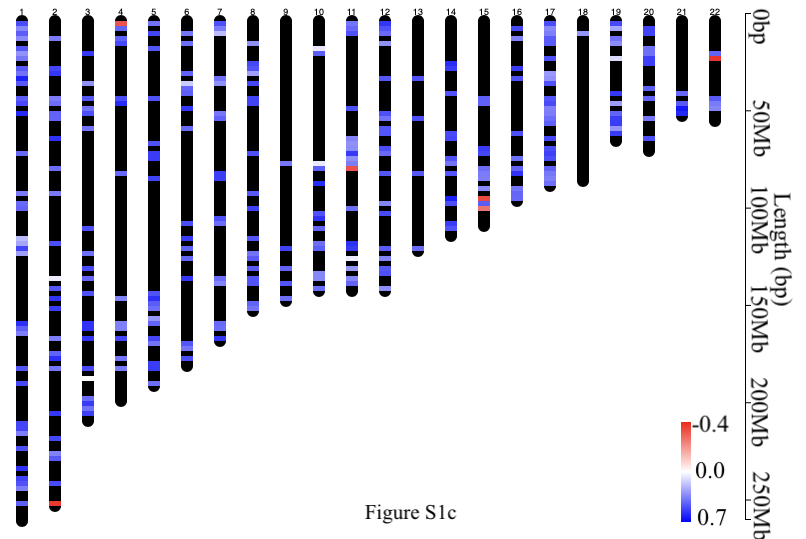

### S2a

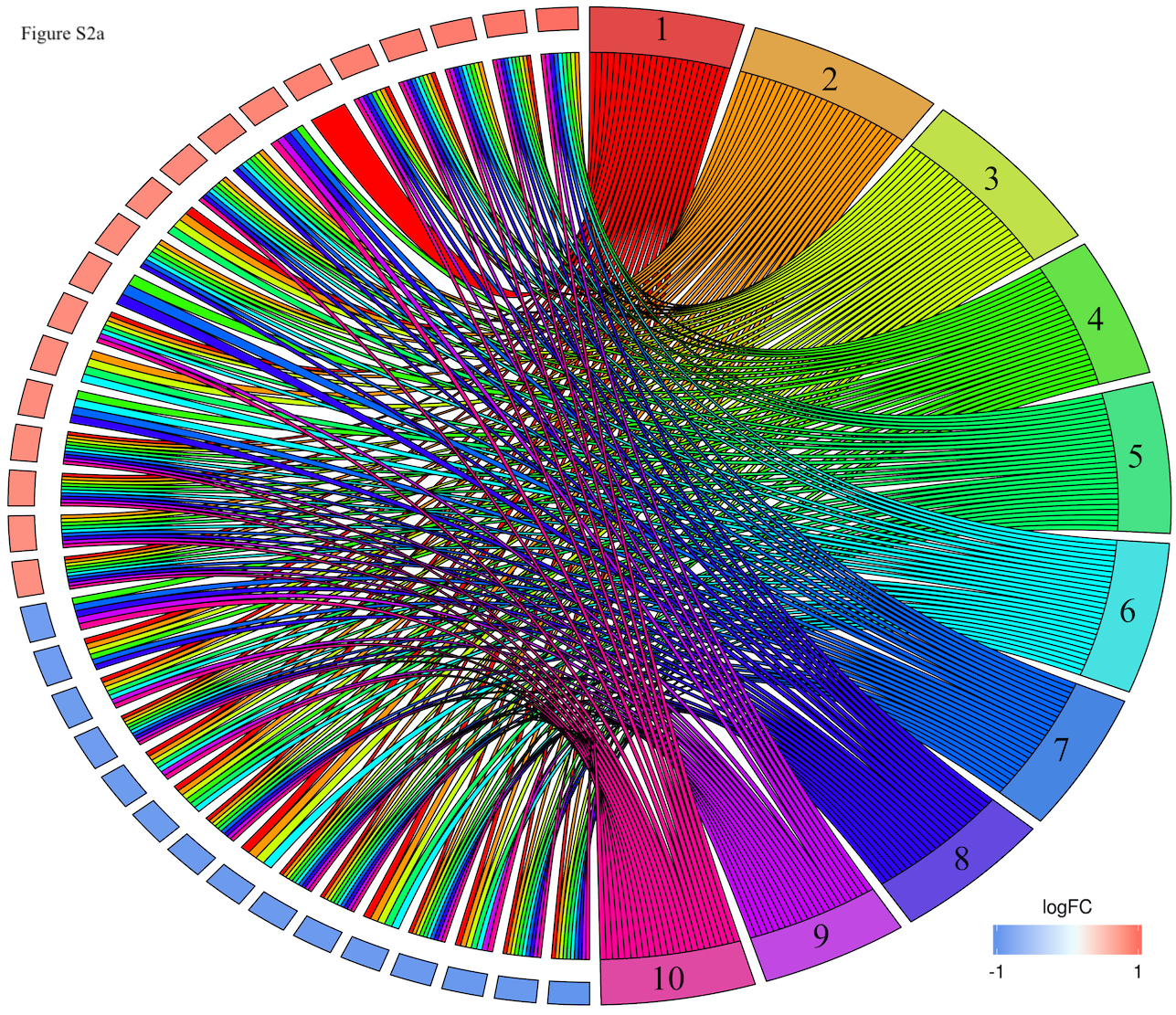

### S2b

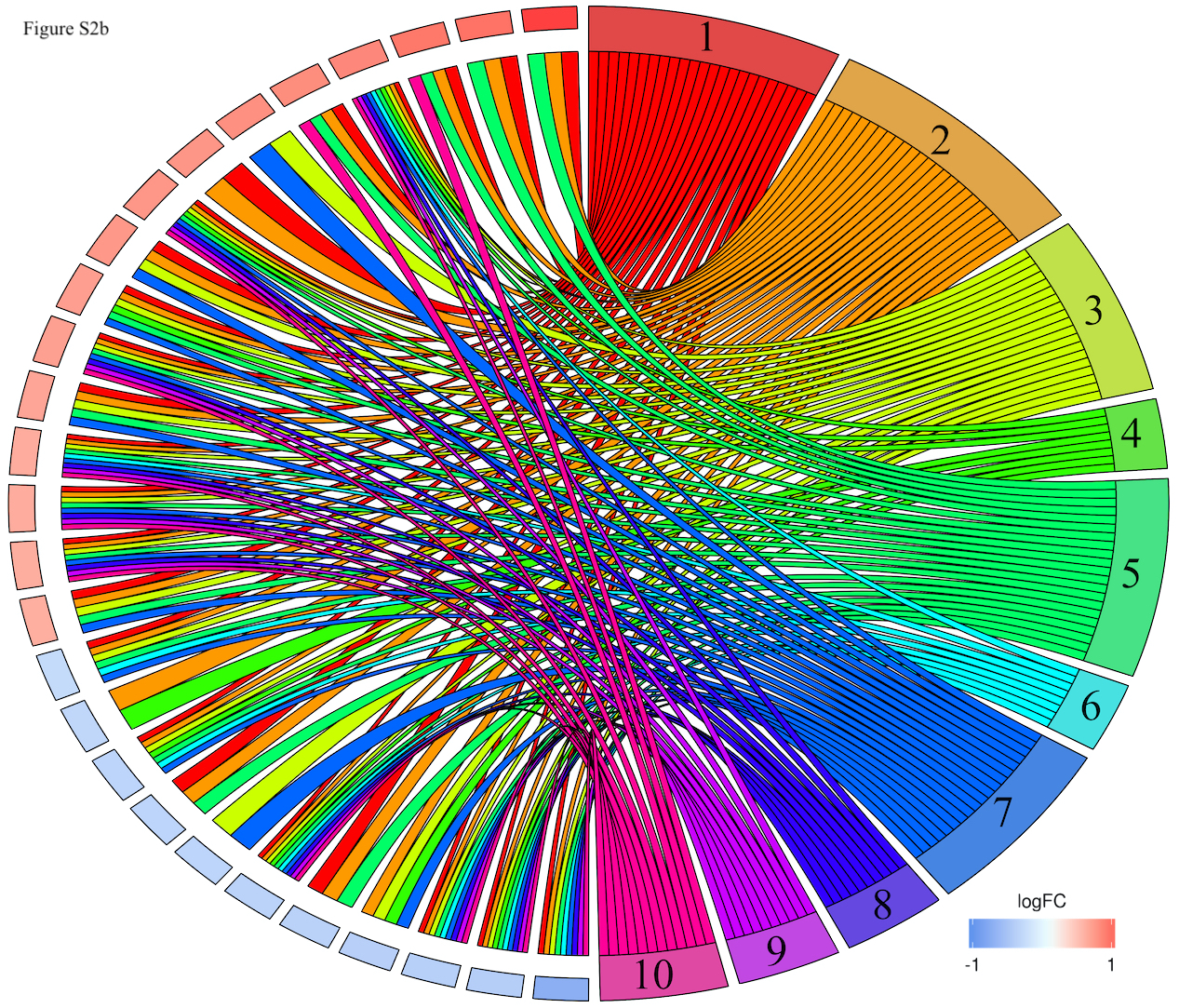

### S2c

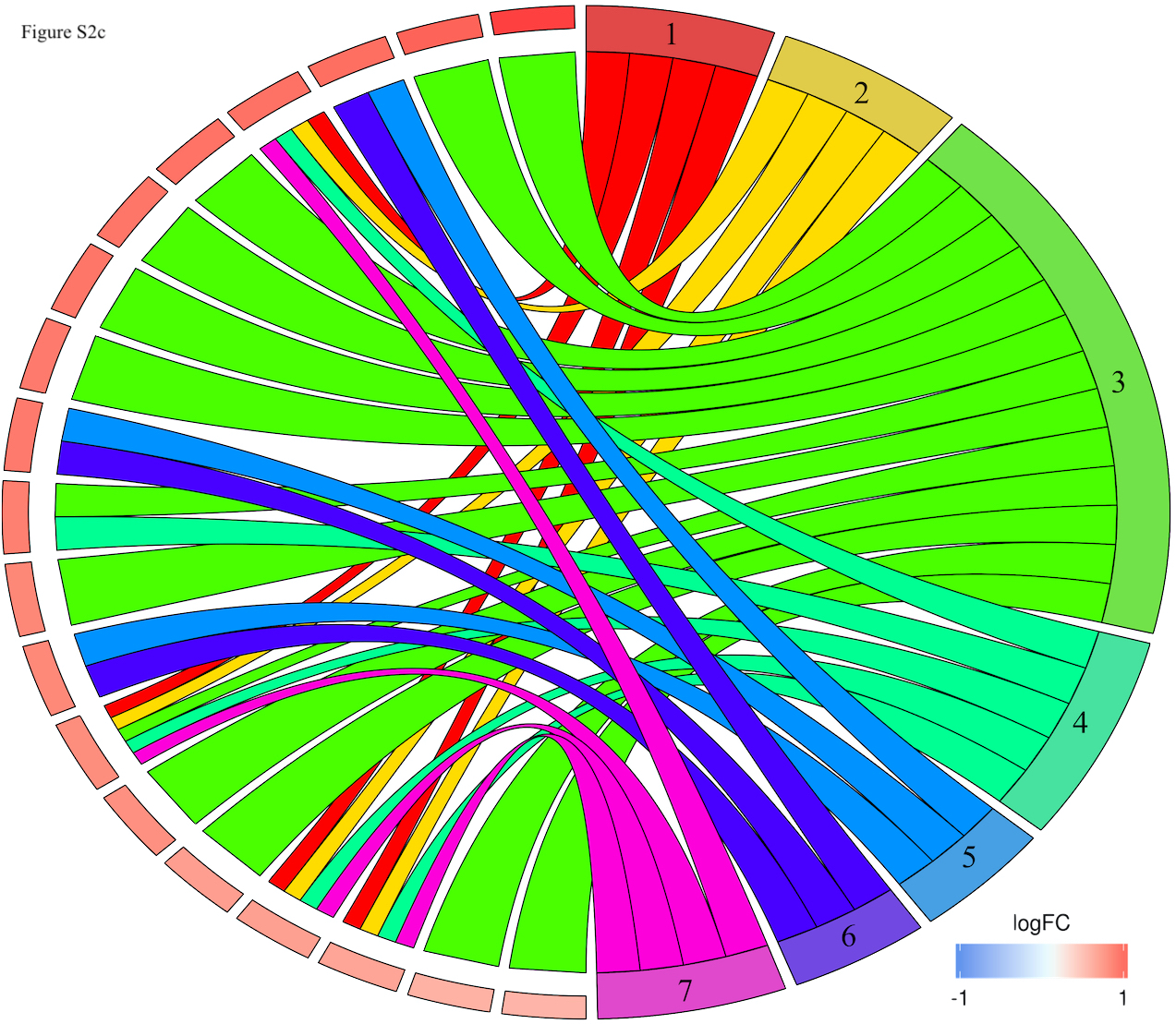

### S2d

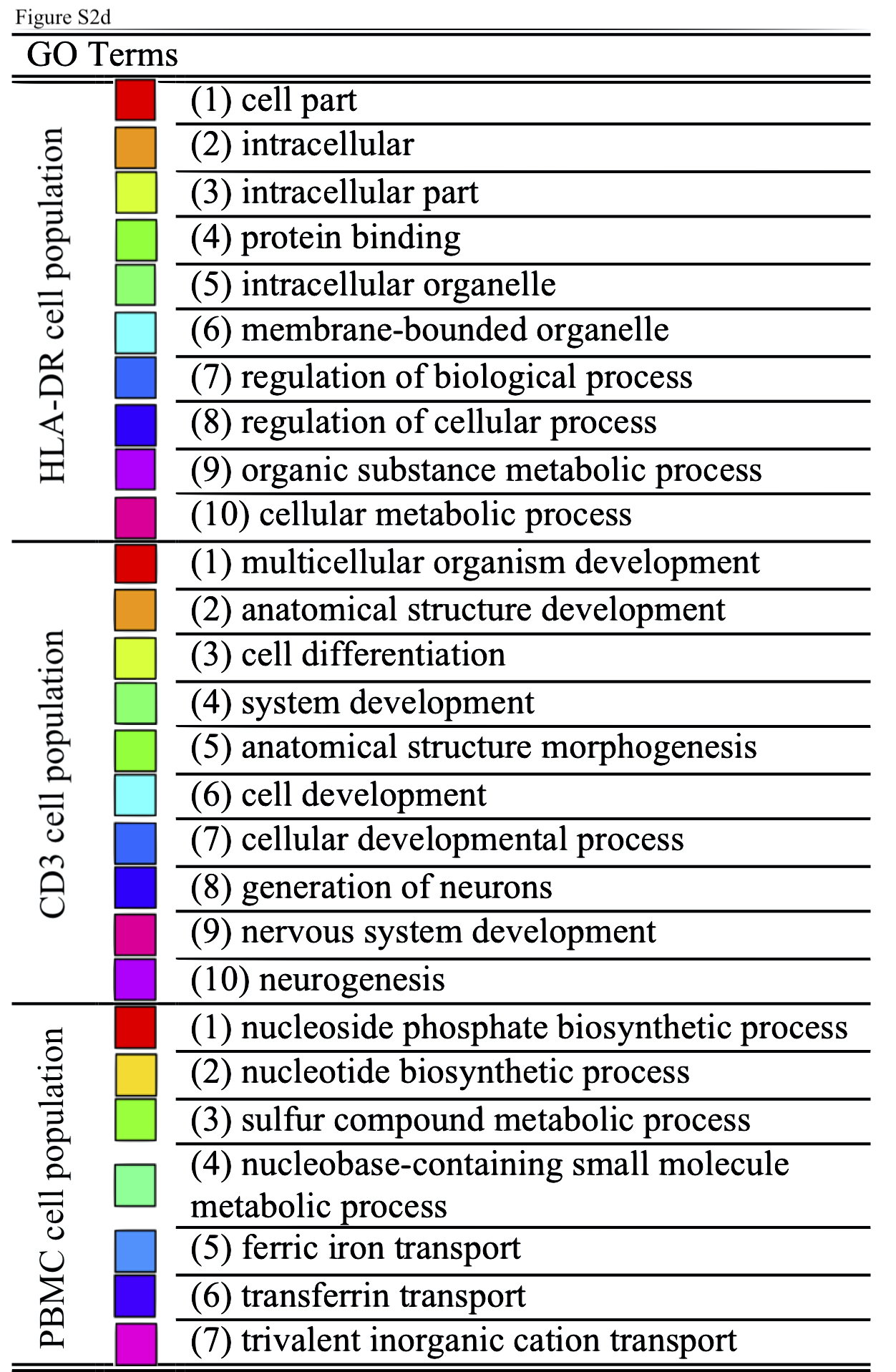
